# Classification accuracy of structural and functional connectomes across different depressive phenotypes

**DOI:** 10.1101/2022.11.22.22282621

**Authors:** Hon Wah Yeung, Aleks Stolicyn, Xueyi Shen, Mark J. Adams, Liana Romaniuk, Gladi Thng, Colin R. Buchanan, Elliot M. Tucker-Drob, Mark E. Bastin, Andrew M. McIntosh, Simon R. Cox, Keith M. Smith, Heather C. Whalley

**Author notes:** These authors share joint senior authorship (Simon R. Cox, Keith M. Smith, Heather C. Whalley).

## Abstract

Phenotyping of major depressive disorder (MDD) in research can vary from study to study, which, together with heterogeneity of the disorder, may contribute to the inconsistent associations with various risk factors including neuroimaging features. These aspects also potentially underlie previous problems with machine learning methods using imaging data to inform predictive biomarkers. In this study we therefore aimed to examine the classification accuracy of structural and functional connectomes across different depressive phenotypes, including separating MDD subgroups into those with and without early childhood adversity (one of the largest risk factors for MDD associated with brain development). We applied logistic ridge regression to classify control and MDD participants defined according to six different MDD definitions in a large community-based sample (*N* = 14, 507). We used brain connectomic data based on six structural and two functional network weightings and conducted a comprehensive analysis to (i) explore how well different connectome modalities predict different MDD phenotypes commonly used in research, (ii) investigate whether stratification of MDD based on the presence or absence of early childhood adversity (measured with the childhood trauma questionnaire) can improve prediction accuracies, and (iii) identify important predictive features that are consistent across MDD phenotypes. We find that functional connectomes consistently outperform structural connectomes as features for MDD classification across phenotypes. Highest accuracy of 61.06% (chance level 50.0%) was achieved when predicting the Currently Depressed phenotype (i.e. the phenotype defined by the presence of more than five symptoms of depression in the past two weeks) with features based on partial correlation functional connectomes. Accuracy of classifying Currently Depressed participants with added CTQ threshold criterion rose to 65.74%. Application of the Jaccard index to assess predictive feature overlap indicated that there were neurobiological differences between MDD patients with and without childhood adversity. Further to that, analysis of predictive features for different MDD phenotypes with binomial tests revealed sensorimotor and visual functional subnetworks as consistently important for prediction. Our results provide the basis for future research, and indicate that differences in sensorimotor and visual subnetworks may serve as important biomarkers of MDD.

## Introduction

Major depressive disorder (MDD) is a disabling psychiatric condition which affects a substantial proportion of the general population around the world. In the clinical setting, diagnosis of MDD relies on subjective reporting of symptoms which can vary between individuals within the same diagnosis. This is further complicated in the research setting where different assessment tools are often used from one research study to another, and this makes the diagnosis of MDD problematic. Different researchers use these different definitions of MDD for their neuroimaging analyses, which likely exacerbates inconsistencies in findings and might underlie problems with using imaging data to classify MDD accurately. Recently, the use of mental health questionnaire-based items to efficiently categorise mood disorder allows researchers to thoroughly explore how depression phenotypes defined with different methods are associated with environmental risk factors, genetics and neuroimaging measures. The large samples now available, together with increased computational capabilities and machine learning techniques, could be key to improving classification and are a step towards understanding depression heterogeneity and potential subtyping.

In the context of Genome Wide Association analyses, Howard et al. (2018) investigated three definitions of MDD - broad depression, probable MDD and International Classification of Diseases based MDD - in UK Biobank (UKB) sample. They reported high genetic correlations among the three MDD phenotypes, indicating a potential core genetic component shared across the MDD definitions. On the other hand, they also found genetic associations that were specific to each MDD phenotype (Howard et al., 2018). Cai et al. (2020) presented five definitions of MDD in the same sample; 3 of the definitions were categorised as minimally phenotyped, and the other two were termed as strictly-defined according to clinical diagnostic criteria (Cai et al., 2020). The authors showed through clustering analysis that the broader definitions exhibited distinct patterns of associations with environmental risk factors as compared to more strictly defined MDD. They also found that the stricter definitions of MDD exhibited higher heritability estimates, more specific genetic architecture, and differing patterns of genetic associations with the strictly defined phenotypes. In terms of analyses of functional brain imaging data, some studies have indicated that associations may be specific to different depressive symptoms. Different depressive symptoms - for example, rumination (Kühn et al., 2012; Wu et al., 2015; Zhu et al., 2012), helplessness and hopelessness (Peng et al., 2014; Yao et al., 2009), and suicidal tendencies (Fan et al., 2013; Zhang et al., 2016) - were found to be associated with abnormal functional connectivity of different brain regions (Brakowski et al., 2017). In structural brain imaging, MDD-related differences were found to be more consistent across depression phenotypes. In our previous work, Harris et al. (2022) studied the structural brain differences, measures for the whole brain (cortical thickness, cortical volume and subcortical volume), cortical lobes and white matter tract types (fractional anisotropy and mean diffusivity), for three main MDD definitions from self-report to clinical definitions in *N* = 39, 300 UKB imaging participants (Harris et al., 2022). They found that the associations with white matter integrity were consistent across depression phenotypes and that phenotype-specific differences were more prominent in cortical thickness measures, despite small overall effect sizes. These studies primarily investigated the influence of phenotyping methods using univariate approaches. In contrast, multivariate analyses and machine learning (ML) approaches combine information from different features for diagnostic classification. There has so far been no large-scale study investigating the effect of depression phenotyping on the results of ML predictive modelling based on structural and functional connectivity data. We hypothesised that our investigated ML models would deploy different decision strategies (reflected in model coefficients), and identify different sets of important features for prediction of different depression phenotypes.

In addition to examining the effects of varying depression phenotyping methods, we here also examine the effect of stratifying depression by presence or absence of early life adversity. Previous studies have shown that early life adversity is associated with increased risk of developing psychiatric disorders including depression in adulthood (Kuzminskaite et al., 2021; Mandelli et al., 2015). Levels of childhood adversity have also been associated with subsequent severity of depression and anxiety symptoms (Huh et al., 2017), as well as abnormal brain connectivity in MDD (Grant et al., 2014; Yu et al., 2019). Luo et al. (2022b) investigated functional connectivity differences between healthy controls (*N* = 80) and MDD cases with and without childhood trauma (*N* = 31 and *N* = 30, respectively). Although both MDD groups had similar alterations in functional connectivity compared to controls, the changes appeared more prominent in cases with childhood trauma. Specifically, MDD cases with childhood trauma exhibited a larger decrease in connectivity between the ventral attention network and sensorimotor network (Luo et al., 2022b). Further to that, Coleman et al. (2020) investigated the genetic relationship between trauma exposure and MDD, where the authors looked at trauma both in childhood and adulthood. Their results showed tht MDD with lifetime trauma exposure had a higher gene-based heritability compared to MDD without trauma exposure (Coleman et al., 2020). These results suggest that MDD cases with childhood trauma may represent a more homogeneous subgroup of MDD, which could be more amenable to diagnostic classification using ML algorithms.

In this study, we investigated diagnostic classification of MDD using connectivity data from the UKB and logistic ridge regression model. We tested the effects of MDD phenotyping on model performance and on the estimated model coefficients. We investigated prediction of six different definitions of MDD using two functional connectome measures (partial correlation and full correlation matrices) and six structural connectome measures. For each diagnostic definition, we maintained a 1-to-1 ratio between cases and controls, with case and control participants matched for age, sex and intracranial volume (ICV), to enable objective comparison of classification accuracies across models. To investigate whether higher classification accuracies may be achieved for MDD subgroups defined by childhood trauma, we repeated the above analyses with only selected MDD participants passing CTQ score threshold. Our main aims were i) to compare classification performances between the different MDD definitions and different brain connectivity modalities, ii) to investigate the effect of childhood trauma score thresholding on classification accuracies, iii) to identify the important features for classifying each MDD phenotype, based on model coefficients, and iv) define important brain subnetworks that may be useful for classifying MDD in general (i.e. important subnetwork changes that are common to different MDD phenotypes).

## Materials and Methods

### Materials

Participants were recruited and brain imaging was completed as part of the UKB study. The six depression phenotypes were derived based on mental health questionnaire (MHQ) items. Five MHQ items were used to derive the CTQ score.

#### Participants

A subset of the UKB participants underwent brain MRI at the UKB imaging centre in Cheadle, Manchester, UK and in Newcastle, UK. The study was approved by the National Health Service Research Ethics Service (No. 11/NW/0382) and by the UKB Access Committee (Project No. 4844 and No. 10279). Written consent was obtained from all participants.

#### Structural Networks

At the time of processing, *N* = 9, 858 participants with compatible T1-weighted and dMRI data were available from the UKB, and the structural connectomes for these participants were derived locally. A full description of structural connectome processing can be found in Buchanan et al. (2020) and Yeung et al. (2022). These processes are described briefly below.

All imaging data were acquired using a single Siemens Skyra 3T scanner (Siemens Medical Solutions, Erlangen, Germany; see http://biobank.ctsu.ox.ac.uk/crystal/refer.cgi?id=2367). Details of the MRI protocol and preprocessing are freely available (Alfaro-Almagro et al., 2018; Miller et al., 2016). Each T1-weighted volume was parcellated into 85 distinct neuroanatomical Regions-Of-Interest (ROI) with FreeSurfer v5.3.0 and 34 cortical structures per hemisphere were identified according to the Desikan-Killany atlas (Desikan et al., 2006). Brain stem, accumbens area, amygdala, caudate nucleus, hippocampus, pallidum, putamen, thalamus and ventral diencephalon were also extracted with FreeSurfer. Whole-brain tractography was performed using an established probabilistic algorithm (BEDPOSTX/ProbtrackX; (Behrens et al., 2007, 2003)) using criteria as described previously (Buchanan et al., 2020). Water diffusion parameters were estimated for FA, which measures the degree of anisotropic water molecule diffusion, and for MD, which measures the magnitude of diffusion. The parameters obtained from NODDI were: ICVF which measures neurite density; ISOVF which measures extracellular water diffusion; and OD which measures the degree of fanning or angular variation in neurite orientation (Zhang et al., 2012). After aligning ROIs from T1-weighted to diffusion space, networks were then constructed by identifying pairwise connections between the 85 ROIs and represented in the form of person-specific 85 *×* 85 adjacency matrices. Six network weightings were computed. Streamline count (SC) was computed by recording the total streamline count (uncorrected) between each pair of ROIs. In addition, five further network weightings (FA, MD, ICVF, ISOVF and OD) were computed by recording the mean value of the diffusion parameter in voxels identified along all interconnecting streamlines between each pair of ROIs.

In total, 8, 183 participants (45.1–78.5 years of age, 3,869 male) remained after participants were excluded following local quality checking or due to failure in processing. Proportional-thresholding was used to keep only connections present in at least 2/3 of subjects (Buchanan et al., 2020; de Reus and van den Heuvel, 2013). 6, 247 out of 8, 183 participants have completed the MHQ questionnaire.

#### Resting-state Functional Networks

*N* = 19, 831 participants underwent a resting state functional MRI (rs-fMRI) assessment and passed the quality check by the UKB. Out of 19, 831 participants, 14, 507 had completed the MHQ questionnaire.

The functional connectome matrices were derived by the UKB imaging project team. The detailed methods of the imaging processing for UKB can be found in previous protocol articles (Alfaro-Almagro et al., 2018; Miller et al., 2016). The processes are described briefly below.

From raw rs-fMRI scans to the correlation and partial correlation matrices, the data went through steps of data preprocessing, parcellation using group independent component analysis (ICA), and connectivity estimation with FSL packages (http://biobank.ctsu.ox.ac.uk/crystal/refer.cgi?id=1977) by the UKB imaging team. The preprocessing steps were completed in the following sequence: motion correction, grand mean intensity normalisation, high-pass temporal filtering, echo-planar image unwarping, gradient distortion correction unwarping, and finally removal of structured artifacts (Miller et al., 2016). A group level ICA was carried out on the first 4,100 participants and the spatial ICA mask was applied to the fMRI scans, parcellating the brain into 100 components. 45 of the 100 components were identified as noise components and were removed. The time-series data for remaining nodes was then used to calculate functional connectivity between node pairs. The full correlation matrices were computed using normalized temporal correlation of the time-series between each pair of nodes. As for the partial correlation matrices, these were computed using partial Pearson correlation with an L2 regularisation applied (rho set as 0.5 for Ridge Regression in FSLNets). All r-scores were then Fisher-transformed into z-scores. This resulted in two 55×55 correlation matrices (correlation and partial correlation) of functional connectivity for each participant. The list of good nodes can be found in: http://https://www.fmrib.ox.ac.uk/datasets/ukbiobank/ group_means/edge_list_d100.txt. An interactive website displaying group-mean maps for each component can be found in: http://www.fmrib.ox.ac.uk/datasets/ukbiobank/group_means/rfMRI_ICA_d100_good_nodes.html. A connectome map of the nodes can be found in: http://www.fmrib.ox.ac.uk/datasets/ukbiobank/netjs_d100/.

#### Depression Phenotypes

A total of 157, 357 participants from the UKB completed an online Mental Health Questionnaire (MHQ) including self-report, clinical lifetime disorder status and experiences of psychiatric symptoms for specific disorders. Davis et al. (2020) presented a detailed implementation for constructing the depression phenotypes from the mental health questionnaire (MHQ) items for UKB (Davis et al., 2020). A number of them have a comparatively loose criteria (which may imply mildly depressed) compared with the others. Some of the depression definitions have a large portion of participant overlap. Here the overlap refers to participants satisfying case criteria for more than one MDD phenotype. We decided to choose one representative (Ever depressed depression phenotype) for the mildly depressed definitions. For the moderate-to-severe MDD phenotypes with large portion of pariticpant overlap, we chose the more severe definitions (Depression Medicated, Ever Severely Depressed, Currently Depressed and Recurrent Depression). Participants who have taken antidepressants were defined as cases for the Depression Medicated phenotype. Ever Depressed was based on Composite International Diagnostic Interview (CIDI) diagnostic criteria. Cases of Ever Severely Depressed had a CIDI severity score of 8. Currently Depressed cases were those who satisfied the criteria for Ever Depressed and also reported current symptoms. Cases of Recurrent Depression had experienced more than one depressive episodes. We also included another MDD definition, Probable Moderate/Severe Depression, described in Smith et al. (2013). In total, we investigated 6 different depression phenotypes and the detailed descriptions of criteria are in Supplementary Material A.1.

#### Childhood adversity

The CTQ score is derived from five childhood trauma-related questionnaire items, which is the same set of items used in (Davis et al., 2020). The item description and scoring system are in Supplementary Material A.2. Based on research indicating that an aggregated trauma score may provide a better marker of risk for adverse outcomes than individual items (Hughes et al., 2017), we took the average score from the five CTQ items as the final CTQ score, which was similar to Warrier and Baron-Cohen (2021). We then investigated the effect of CTQ score thresholding on model performance and model coefficients.

Four different CTQ score thresholds (None, 0.2, 0.4 and 0.6) corresponding to the 50th, 70th and 80th quantile, were applied (i.e., MDD cases falling below the CTQ threshold were excluded from the subgroup). The use of different thresholds can help verify the relationship between CTQ severity cut-off and classification accuracies. Table 1 shows the sample size of MDD cases for each definition and for each CTQ score threshold.

**Table 1.**
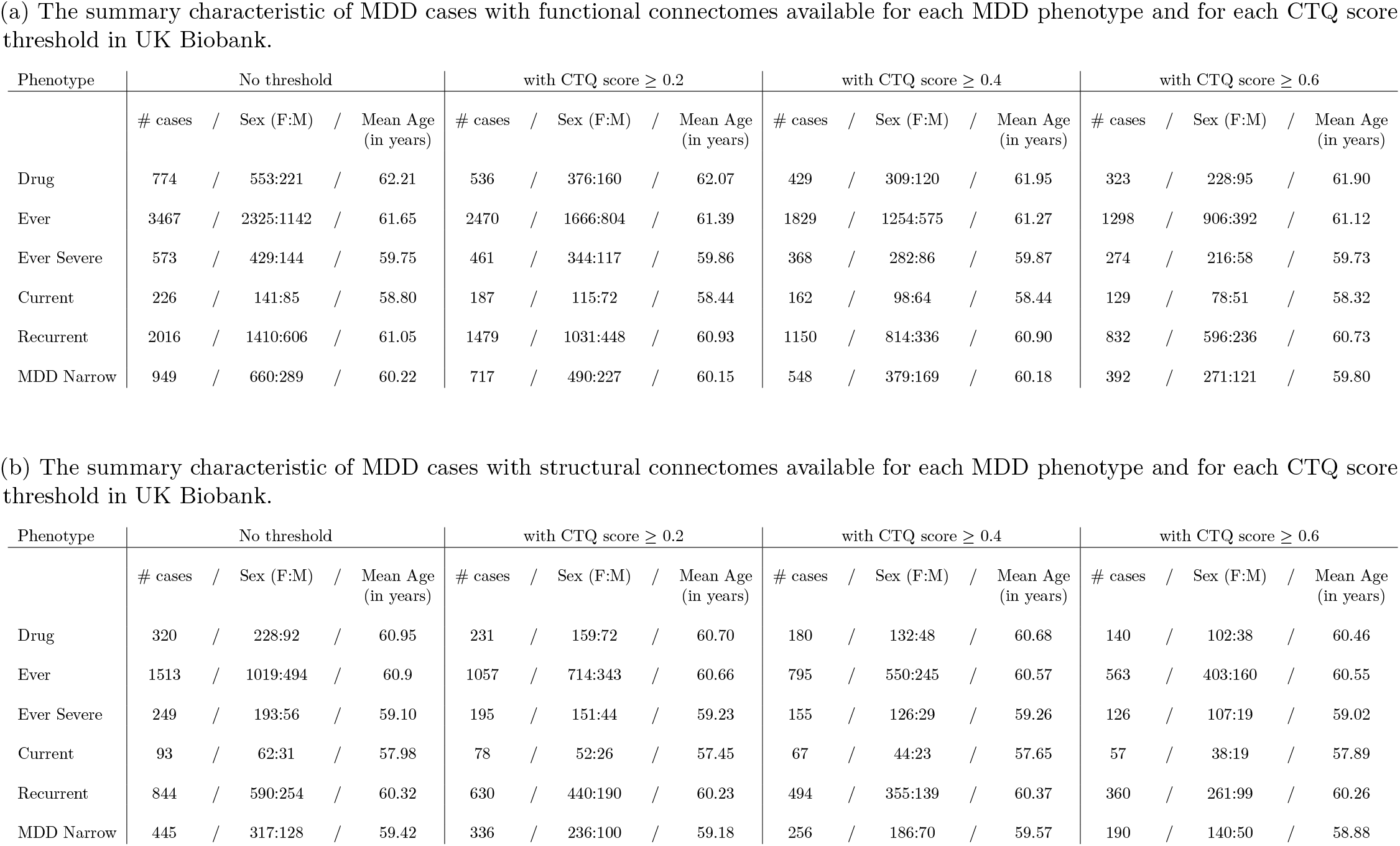
The summary characteristic of MDD cases for each MDD phenotype and for each CTQ score threshold in UK Biobank. Drug = Depression Medicated, Ever = Ever Depressed, Ever Severe = Ever Severely Depressed, Current = Currently Depressed, Recurrent = Recurrent Depression without Bipolar Disorder, MDD Narrow = Probable Moderate/Severe Depression

### Methods

#### Classification model

Previous neuroimaging ML studies indicate that classical ML models achieved accuracy comparable to Deep Learning (DL) models (He et al., 2020). Moreover, work by Schulz et al. (2020) indicate that simple linear models were just as competitive as non-linear models, sometimes even outperforming non-linear models, in predicting common phenotypes from brain scans (Schulz et al., 2020). We have found similar results in a previous study of structural connectomes that ridge regression tends to be more consistent than other models in terms of model coefficients (Yeung et al., 2022). Therefore, a logistic ridge regression model was chosen for classification modelling in the current study.

#### Feature Inputs

In this study, the inputs to the classification model are the vectorised upper triangular non-zero entries of the connectivity matrices (Number of features: functional connectivity, *N* = 1, 485; structural connectivity, *N* = 2, 210). We additionally examined combined inputs of functional and structural connectivity to the models, where one of the six structural modalities is stacked on one of the two functional modalities to give a long vector of connectivity features (Number of features, *N* = 3, 695). The combinations give 12 sets of combined connectivity features.

#### Regularization parameter Optimization

In order to find the optimal regularisation parameter, *λ*, we tested different values ranging from 1 *×* 10^*−*3^*to*1 *×* 10^3^. The *λ* that optimized validation accuracies at each iteration of the inner loop was chosen, so the *λ*s are different for each fold, each MDD definition and each connectivity modality.

#### Case-control matching and Correction for confounders

We aimed to run classification models with a 1-to-1 ratio between cases and controls for each of the six MDD phenotypes. Those who are not defined as cases in any of the 17 MDD definitions (derived from Davis et al. (2020) and Smith et al. (2013)) formed the full healthy control sample (Full-HC). For each case participant from the six MDD-phenotype samples, we selected a control from the Full-HC matched by sex, with smallest difference in age and in ICV. In the case with CTQ thresholding, the CTQ criterion was applied to the MDD samples first before case-control matching so that the classification models always run on a sample with 1-to-1 ratio between MDD cases and healthy controls.

Following the above stringent criteria (particularly with minimal differences in ICV), we confirmed that there was exactly one matched control for each case, and therefore none of the control samples were drawn at random. Participants with other major neurological or psychiatric disorders (namely schizophrenia, bipolar, multiple personality disorder, autism, intellectual disability, Parkinson’s disease, multiple sclerosis or cognitive impairment) were excluded.

#### Experimental setup

We ran a nested cross-validation (CV) to evaluate the performance. In the nested 5-fold CV, the training data (5/6 folds) of each of the six iterations of the outer 6-fold cross-validation was split into 5 folds. Connectome features were *z*-normalized based on training data. The optimal model, which achieved highest validation accuracy, was chosen in each of the inner iteration. Therefore, this amounts to a total of 30 evaluations of classification accuracies. The split was carefully done so that the cases and control maintained a 1-to-1 ratio with age, sex and ICV matched for any of the training, validation and test set. Test accuracies were averaged across the folds to get performance estimates.

#### Identifying important features

Model coefficients tell us about feature importance in classification tasks and help us identify the features associated with the specific MDD definition of interest. A simple way to compare feature importance is to observe the magnitudes of the beta coefficients. However, beta coefficients were data-specific and the ranking by beta coefficients may not be generalisable to new data.

In order to identify the features that were truly important, we believed that there were 2 necessary conditions:

1. The feature should be consistently in the top 50%, by coefficient magnitudes, in all of the 30 models trained in the nested CV.
2. The feature should always have the same coefficient sign in all of the 30 models trained in the nested CV.

#### Identifying important subnetworks

The majority of biological findings in MDD are of small effects and we claimed that this implied the uncertainty in ML model coefficients’ magnitudes for MDD. Therefore, in this study we compared the number of important features (as defined above) within a subnetwork (or between two subnetworks) to the number expected by random chance using a binomial test. The binomial test offers a more objective measure to assess the importance of a functional subnetwork than a simple aggregation of model coefficients. The binomial test is used for testing hypotheses about probability of success, which is therefore appropriate for checking whether there are significantly more/less important features being distributed to certain subnetworks in the connectomes. Let *N* be the number of nodes in the connectome, *S*_*i*_ be the size of subnetwork *i, K* be the total number of important features identified for a MDD phenotype and *O* be the matrix representing the number of important features between/within subnetwork. Then, the matrix *P* representing the probability of having an edge between subnetwork *i* and *j* or within a subnetwork *i* is given by

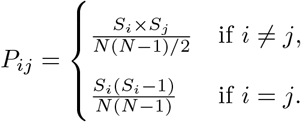

and the *p*-value matrix, *B*, from the binomial test is given by,

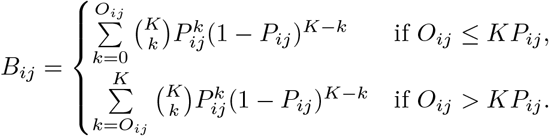

## Results

### Depression classification performances based on different connectome modalities without CTQ threshold

We applied logistic ridge regression based on structural and functional imaging features to classify the six MDD phenotypes identified above. Figure 1 shows the classification results for different MDD phenotypes based on different connectome modalities and the exact numbers are shown in Supplementary Materials B, Table 2(a) - Table 2(h). The chance-level accuracy of all the classifiers below is 50%.

**Table 2.** Number of important features identified by the models for different depression phenotypes (with and without CTQ threshold at 0.4) based on different connectome modalities. Drug = Depression Medicated, Ever = Ever Depressed, Ever Severe = Ever Severely Depressed, Current = Currently Depressed, Recurrent = Recurrent Depression without Bipolar Disorder, MDD Narrow = Probable Moderate/Severe Depression

| MDD phenotype | Drug | Ever | Ever severe | Current | Recurrent | MDD narrow |
| --- | --- | --- | --- | --- | --- | --- |
| # Important Feature (without CTQ threshold) | 41 | 76 | 43 | 60 | 31 | 43 |
| # Important Feature (with CTQ threshold 0.4) | 48 | 80 | 48 | 65 | 30 | 57 |
| Number of Overlap | 15 | 33 | 12 | 27 | 10 | 15 |
| Jaccard index | 0.2027 | 0.2683 | 0.1519 | 0.2755 | 0.1961 | 0.1765 |

(a) CORRELATION FUNCTIONAL CONNECTOME
| MDD phenotype | Drug | Ever | Ever severe | Current | Recurrent | MDD narrow |
| --- | --- | --- | --- | --- | --- | --- |
| # Important Feature (without CTQ threshold) | 60 | 86 | 49 | 84 | 87 | 53 |
| # Important Feature (with CTQ threshold 0.4) | 69 | 27 | 59 | 88 | 57 | 47 |
| Number of Overlap | 22 | 10 | 23 | 41 | 18 | 14 |
| Jaccard index | 0.2056 | 0.0971 | 0.2706 | 0.3130 | 0.1429 | 0.1628 |

(b) PARTIAL CORRELATIONS FUNCTIONAL CONNECTOMES
| MDD phenotype | Drug | Ever | Ever severe | Current | Recurrent | MDD narrow |
| --- | --- | --- | --- | --- | --- | --- |
| # Important Feature (without CTQ threshold) | 87 | 45 | 71 | 81 | 65 | 86 |
| # Important Feature (with CTQ threshold 0.4) | 106 | 70 | 82 | 43 | 90 | 107 |
| Number of Overlap | 33 | 17 | 30 | 31 | 27 | 28 |
| Jaccard index | 0.2063 | 0.1735 | 0.2439 | 0.3333 | 0.2109 | 0.1697 |

(c) MEAN DIFFUSIVITY STRUCTURAL CONNECTOME
| MDD phenotype | Drug | Ever | Ever severe | Current | Recurrent | MDD narrow |
| --- | --- | --- | --- | --- | --- | --- |
| # Important Feature (without CTQ threshold) | 102 | 31 | 80 | 85 | 56 | 80 |
| # Important Feature (with CTQ threshold 0.4) | 87 | 79 | 98 | 130 | 83 | 93 |
| Number of Overlap | 28 | 12 | 35 | 45 | 22 | 28 |
| Jaccard index | 0.1739 | 0.1224 | 0.2448 | 0.2647 | 0.1880 | 0.1931 |

Table 2: Number of important features identified by the models for different depression phenotypes (with and without CTQ threshold at 0.4) based on different connectome modalities.
| MDD phenotype | Drug | Ever | Ever severe | Current | Recurrent | MDD narrow |
| --- | --- | --- | --- | --- | --- | --- |
| # Important Feature (without CTQ threshold) | 70 | 39 | 66 | 74 | 40 | 65 |
| # Important Feature (with CTQ threshold 0.4) | 67 | 37 | 101 | 105 | 54 | 73 |
| Number of Overlap | 24 | 8 | 34 | 38 | 13 | 21 |
| Jaccard index | 0.2124 | 0.1176 | 0.2556 | 0.2695 | 0.1605 | 0.1795 |

(e) STREAMLINE COUNT STRUCTURAL CONNECTOME
| MDD phenotype | Drug | Ever | Ever severe | Current | Recurrent | MDD narrow |
| --- | --- | --- | --- | --- | --- | --- |
| # Important Feature (without CTQ threshold) | 108 | 56 | 81 | 76 | 50 | 73 |
| # Important Feature (with CTQ threshold 0.4) | 82 | 61 | 90 | 114 | 75 | 91 |
| Number of Overlap | 37 | 13 | 26 | 43 | 18 | 26 |
| Jaccard index | 0.2418 | 0.1250 | 0.1793 | 0.2925 | 0.1682 | 0.1884 |

(f) ORIENTATION DISPERSION STRUCTURAL CONNECTOME
| MDD phenotype | Drug | Ever | Ever severe | Current | Recurrent | MDD narrow |
| --- | --- | --- | --- | --- | --- | --- |
| # Important Feature (without CTQ threshold) | 81 | 48 | 64 | 98 | 58 | 69 |
| # Important Feature (with CTQ threshold 0.4) | 89 | 66 | 90 | 109 | 52 | 73 |
| Number of Overlap | 26 | 14 | 23 | 43 | 12 | 23 |
| Jaccard index | 0.1806 | 0.1400 | 0.1756 | 0.2622 | 0.1224 | 0.1933 |

(g) ISOTROPIC VOLUME FRACTION STRUCTURAL CONNECTOME
| MDD phenotype | Drug | Ever | Ever severe | Current | Recurrent | MDD narrow |
| --- | --- | --- | --- | --- | --- | --- |
| # Important Feature (without CTQ threshold) | 83 | 66 | 93 | 104 | 74 | 98 |
| # Important Feature (with CTQ threshold 0.4) | 94 | 83 | 118 | 79 | 112 | 79 |
| Number of Overlap | 37 | 17 | 39 | 44 | 35 | 23 |
| Jaccard index | 0.2643 | 0.1288 | 0.2267 | 0.3165 | 0.2318 | 0.1494 |

(h) INTRA-CELLULAR VOLUME FRACTION STRUCTURAL CONNECTOME

**Figure 1.**
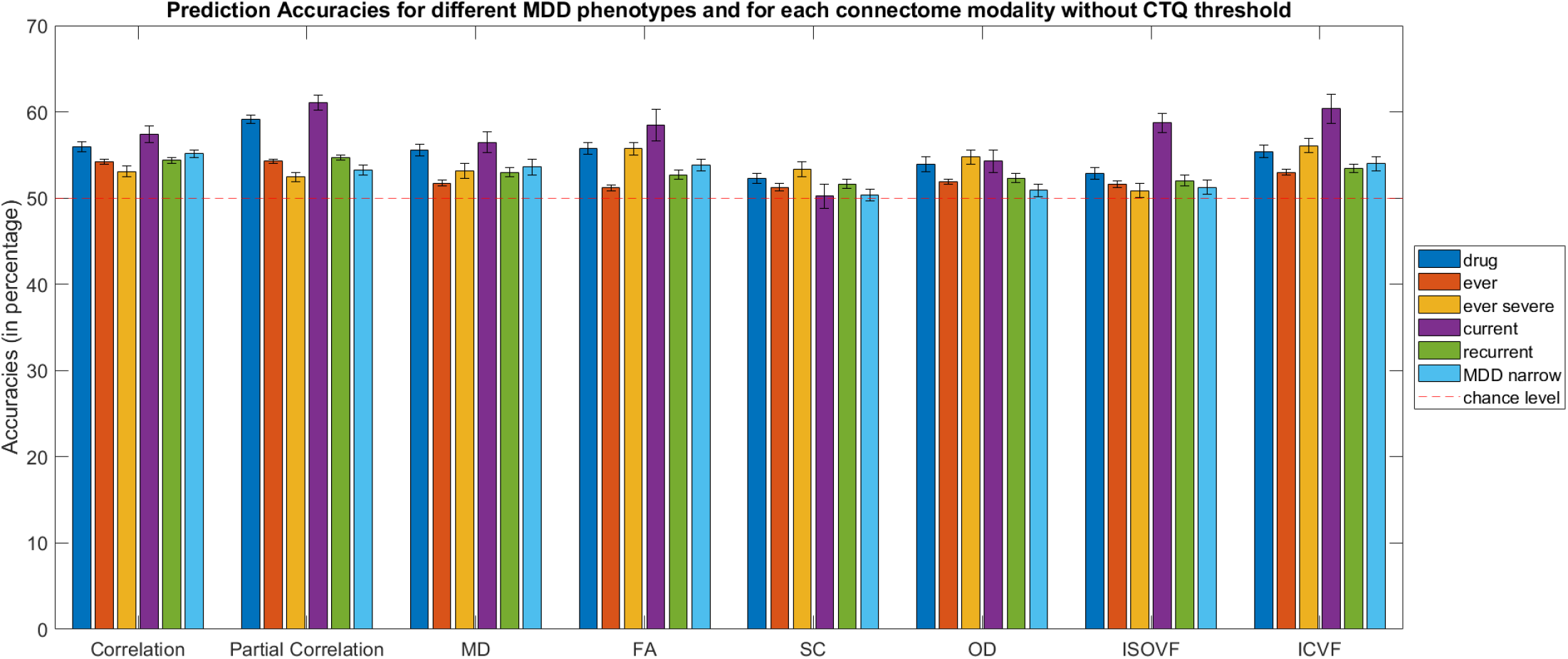
Bar plot for MDD phenotypes (without Childhood Trauma Questionnaire (CTQ) threshold scores) classification accuracies (mean percentage with error bars showing the standard error) with functional connectomes for the test sets. Drug = Depression Medicated, Ever = Ever Depressed, Ever Severe = Ever Severely Depressed, Current = Currently Depressed, Recurrent = Recurrent Depression without Bipolar Disorder, MDD Narrow = Probable Moderate/Severe Depression, Corr = functional correlation connectivity, pCorr = functional partial correlation connectivity, MD = mean diffusivity, FA = fractional anisotropy, SC = streamline count, OD = orientation dispersion, ISOVF = isotropic volume fraction, ICVF = intra-cellular volume fraction

In terms of functional connectomes, for correlation functional connectivity (Corr), the model classifications for five of the MDD definitions had comparable performances, with test accuracies ranging from 53.1% - 56.0%, and slightly better test accuracies for Currently Depressed (57.4%). As for partial correlation functional connectivity (pCorr), the mean test accuracies for Depression Medicated (59.2%) and Currently Depressed (61.1%) outperformed the other definitions. The test accuracies for the other four definitions ranging from 52.4% to 54.7%.

For structural connectomes, the classification accuracies for the test sets (overall range: 50.3—60.4%) were mostly lower than those based on functional connectomes. ICVF gave slightly better overall MDD classification test accuracies (*≥* 52.9%) than the other five network weights (*≥* 50.3%). Similar to findings with functional connectomes, the classification performance for Currently Depressed phenotype was the best among all other phenotypes for all network weights (54.3—60.4%), with the exception of SC (50.3%).

### Depression classification performances for Currently Depressed phenotype based on different connectome modalities with CTQ threshold

It was found that different connectome modalities generally achieved best test accuracies for the Currently Depressed phenotype, and improvements in classifications were mostly found in models with CTQ threshold at 0.4, see Supplementary Materials B, Table 2(a) - (h). Figure 2 shows the test accuracies for Currently Depressed phenotype based on different connectome modalities with and without CTQ threshold.

**Figure 2.**
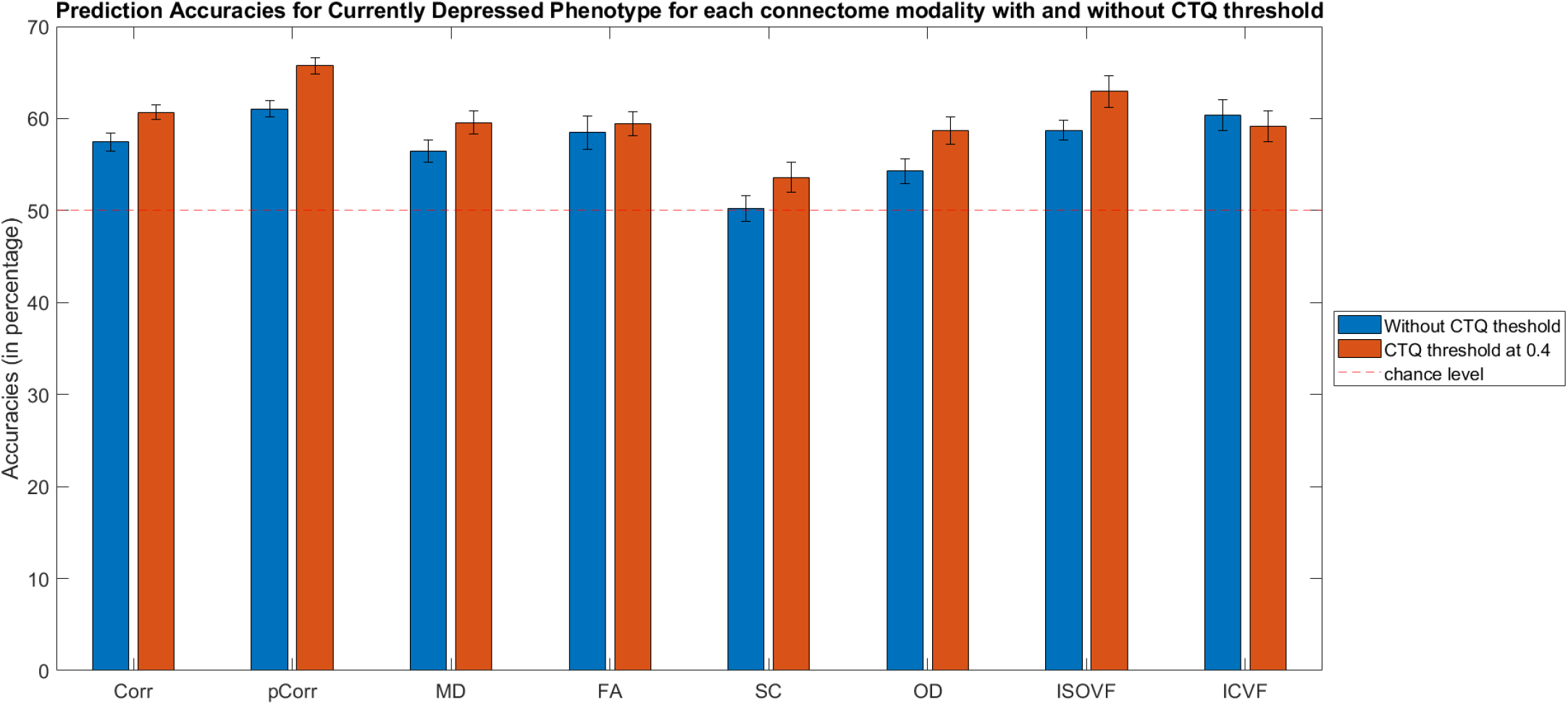
Bar plot for Currently Depressed Phenotype (with and without Childhood Trauma Questionnaire (CTQ) threshold scores) classification accuracies (mean percentage with error bars showing the standard error) with different connectome modalities for the test sets. Corr = functional correlation connectivity, pCorr = functional partial correlation connectivity, MD = mean diffusivity, FA = fractional anisotropy, SC = streamline count, OD = orientation dispersion, ISOVF = isotropic volume fraction, ICVF = intra-cellular volume fraction

With CTQ threshold, the pCorr gave the best test accuracy (65.74%), with an improvement of 4.68% from the model without CTQ threshold. The MDD classification test accuracies based on Corr improved from 57.44% to 60.68%. The model based on ISOVF also saw significant improvement, with test accuracy rising from 58.73% to 62.94%. Higher accuracies were also seen in other modalities except for ICVF.

Furthermore, the more severe phenotype (where there were also fewer number of cases) usually exhibited higher accuracies. By checking over all the depression phenotypes and at different levels of CTQ threshold, we observed negative correlations between test accuracies and sample size, and the correlations were statistically significant (i.e. *p*-value < 0.05) for most of the structural and functional connectivity modalities except for SC, OD and ISOVF (Corr : *r* = *−*0.4542 ; pCorr : *r* = *−*0.4179 ; MD : *r* = *−*0.4848 ; FA : *r* = *−*0.5031 ; SC : *r* = *−*0.0069 ; OD : *r* = *−*0.3463 ; ISOVF : *r* = *−*0.2882 ; ICVF : *r* = *−*0.5713).

### Depression classification performances for Currently Depressed phenotype based on combined connectivity

We further investigated the added value of structural connectivity to functional connectivity in classification modelling of MDD. Figure 3 shows the classification performances for Currently Depressed phenotypes based on the 12 sets of combined connectivity at different CTQ thresholds. The samples here were restricted to the participants with both functional and structural connectivity data available, and therefore were different from the samples used in the above section. Results showed that adding structural connectivity to Corr boosted the test accuracies (improved by 0.9 - 5.9%, averaged across CTQ thresholds), and the improvements were more prominent for ISOVF (+ 5.9%) and ICVF (+ 3.9%). On the other hand, the added value of structural connectivity to pCorr was not clear. Improvement was only seen with ICVF. Due to the differences in nature of the connectivity (i.e. dynamic vs static), functional and structural connectomes were investigated separately in the following.

**Figure 3.**
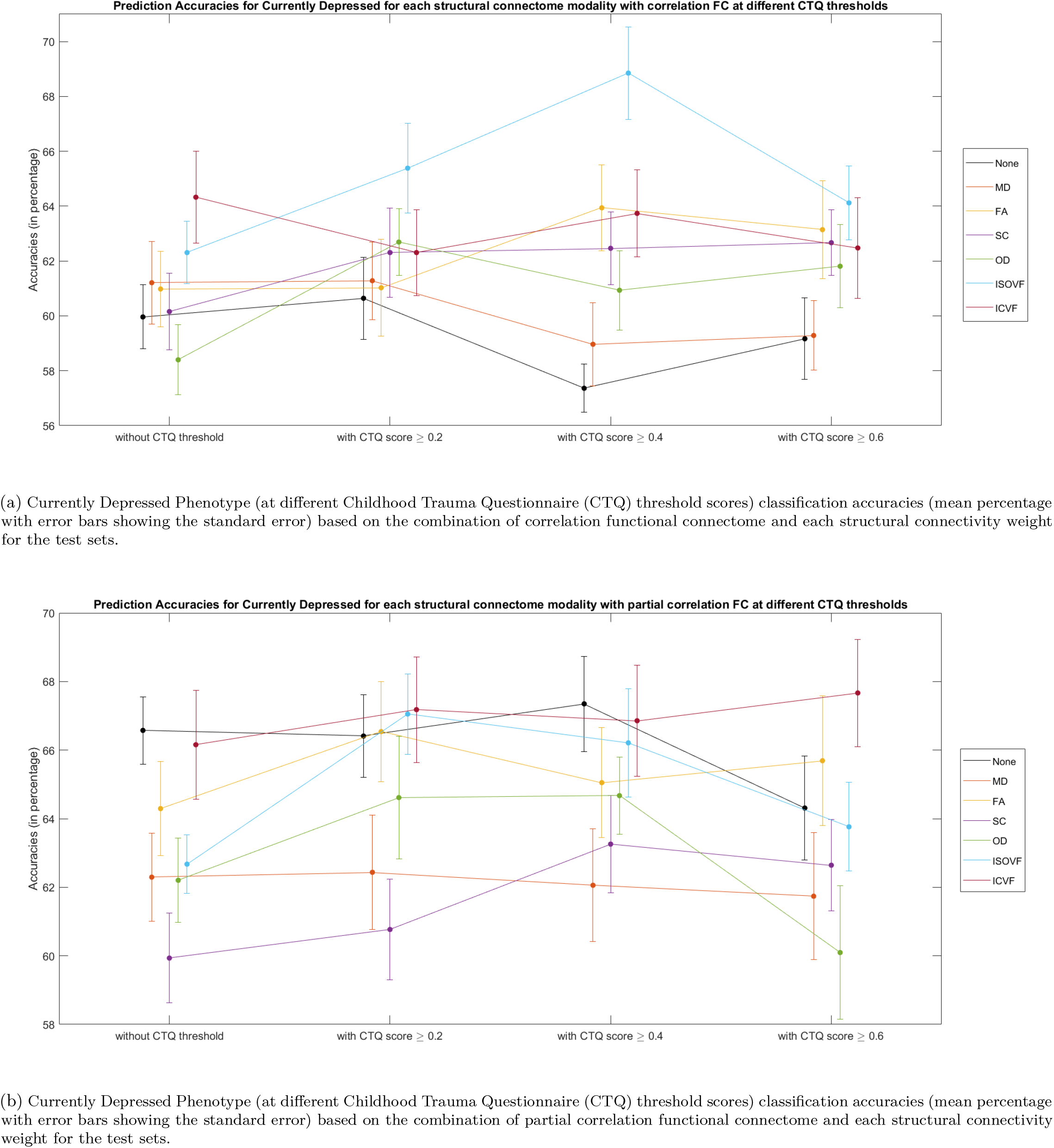
Currently Depressed Phenotype (at different Childhood Trauma Questionnaire (CTQ) threshold scores) classification accuracies (mean percentage with error bars showing the standard error) based on the combination of functional connectome and each structural connectivity weight for the test sets. FC = functional connectome, None = no structural connectivity added to the model, MD = mean diffusivity, FA = fractional anisotropy, SC = streamline count, OD = orientation dispersion, ISOVF = isotropic volume fraction, ICVF = intra-cellular volume fraction. Note that the samples here are restricted to the participants with both functional and structural connectivity data available

### Comparing important edges with and without CTQ threshold

As mentioned in the Methods, we used the two criteria (i.e. feature coefficients consistently with top 50% in magnitude ranking and with same sign across the CV models) to identify important features for the specific MDD phenotype for each imaging modality. In general, we found that the number of important features identified was positively and significantly correlated with models’ test accuracies for most of the functional and structural connectome weights, except for MD, FA and SC (Corr : *r* = 0.5024 ; pCorr : *r* = 0.6053 ; MD : *r* = *−*0.0859 ; FA : *r* = 0.3775 ; SC : *r* = 0.3080 ; OD : *r* = 0.5164 ; ISOVF : *r* = 0.6180 ; ICVF : *r* = 0.4220), and was negatively and significantly correlated with the sample size for most of the functional and structural connectome weights, except for Corr and pCorr (Corr : *r* = *−*0.0937 ; pCorr : *r* = 0.0417 ; MD : *r* = *−*0.4493 ; FA : *r* = *−*0.7883 ; SC : *r* = *−*0.7887 ; OD : *r* = *−*0.7511 ; ISOVF : *r* = *−*0.7913 ; ICVF : *r* = *−*0.5593). Since the CTQ threshold at 0.4 generally performed the best out of the three thresholds, we chose to compare the important features from models without CTQ threshold and models with CTQ threshold at 0.4.

Table 2 shows the number of important edges identified for each of the depression phenotypes and for each connectome modality as well as the number of feature overlaps between models with and without the CTQ threshold. We also presented the Jaccard index which provided a more objective estimate of the degree of overlap that accounts the differences in the number of important features identified by the models. Formulation of the Jaccard index is presented in the Supplementary Materials A.3. We found that all models have less than 10% of the edges being identified as robust and important features for predicting depression phenotypes. Moreover, the set of important features identified in the models without CTQ threshold were mostly different from those identified in the models with CTQ threshold (Jaccard = 0.10 *−* 0.33). A more detailed comparison of the important functional connectome features for the different MDD phenotypes (with and without CTQ threshold) are shown in the following subsection.

### Post-hoc analyses of important edges in functional connectivity

Since the model performance based on functional connectomes were generally better than that based on structural connectomes, the following analysis focuses on functional connectomes. We also built the sex stratified MDD classification models based on functional connectomes. The same experimental setup was used for male and female MDD classification models. Details of the test accuracies are in the Supplementary Materials B.1, Table 3. It was found that the original models performed better than the sex specific ones. Since there was no improvement in test accuracies, we only analysed the original models on the six different MDD phenotypes in the following section, and with more emphasis on the classification models on the currently depressed phenotype, the phenotype with the highest test accuracies. After identifying the important edges, a binomial test was used to determine the importance of certain functional subnetworks in the context of MDD classification modelling. The normalised predictive power for the subnetworks were also reported. Details and formulation of the normalised predictive coefficients are presented in the Supplementary Materials A.4.

**Table 3.** Normalised predictive coefficient between or within subnetworks for the six MDD classification without CTQ threshold based on correlation functional connectomes. The *p*-values from binomial test were in the brackets, red cell with bold text = significantly fewer edges, green cell with bold text = significantly more edges. Sensorimotor network = SMN, visual network = VN, executive control and attention network = EC_AN, cingulo-opercular network = CON, default mode network = DMN and extended default mode network = eDMN

| Functional Subnetwork | SMN | VN | EC_AN | CON | DMN | eDMN |
| --- | --- | --- | --- | --- | --- | --- |
| SMN | **0.0054 (0.0077)** |  |  |  |  |  |
| VN | **0.0045 (0.0485)** | 0 (0.758) |  |  |  |  |
| EC_AN | -0.0163 (0.2771) | -0.0091 (0.068) | 0.008 (0.5746) |  |  |  |
| CON | 0.0051 (0.1004) | 0 (0.2832) | -0.005 (0.4284) | 0 (0.3656) |  |  |
| DMN | **0.0108 (0.0241)** | 0 (0.4331) | 0 (0.0646) | 0.0022 (0.1865) | 0 (0.6595) |  |
| eDMN | **-0.0049 (0.2809)** | **0.0151 (0.0351)** | 0.0038 (0.4284) | -0.0036 (0.3375) | 0 (0.219) | -0.0105 (0.262) |

(a) DEPRESSION MEDICATED
| Functional Subnetwork | SMN | VN | EC_AN | CON | DMN | eDMN |
| --- | --- | --- | --- | --- | --- | --- |
| SMN | -0.0052 (0.081) |  |  |  |  |  |
| VN | -0.0011 (0.2701) | **0.0162 (0.0018)** |  |  |  |  |
| EC_AN | -0.0011 (0.4177) | 0.0062 (0.2257) | 0.0014 (0.4157) |  |  |  |
| CON | -0.0014 (0.3131) | -0.001 (0.1991) | -0.004 (0.1288) | **-0.0067 (0.0375)** |  |  |
| DMN | 0.001 (0.6311) | 0 (0.212) | -0.0026 (0.4504) | 0.0016 (0.2317) | 0 (0.4623) |  |
| eDMN | -0.0026 (0.089) | **-0.0029 (0.0083)** | 0.0079 (0.3156) | -0.0013 (0.0758) | 0.0043 (0.299) | 0.0039 (0.2802) |

(b) EVER DEPRESSED
| Functional Subnetwork | SMN | VN | EC_AN | CON | DMN | eDMN |
| --- | --- | --- | --- | --- | --- | --- |
| SMN | 0 (0.2663) |  |  |  |  |  |
| VN | 0 (0.2293) | **0.0432 (0.0002)** |  |  |  |  |
| EC_AN | 0 (0.1756) | 0.0009 (0.5894) | **0.0116 (0.0204)** |  |  |  |
| CON | -0.0073 (0.5122) | 0.0004 (0.1411) | -0.0054 (0.1996) | 0.0089 (0.72) |  |  |
| DMN | 0 (0.1697) | 0 (0.4158) | -0.013 (0.3018) | 0 (0.2034) | 0.0143 (0.0702) |  |
| eDMN | -0.0006 (0.5122) | -0.0042 (0.6241) | 0.0011 (0.5942) | **0.0073 (0.0283)** | -0.0005 (0.2052) | 0 (0.3481) |

(c) EVER SEVERELY DEPRESSED
| Functional Subnetwork | SMN | VN | EC_AN | CON | DMN | eDMN |
| --- | --- | --- | --- | --- | --- | --- |
| SMN | 0.0274 (0.1087) |  |  |  |  |  |
| VN | -0.0147 (0.0514) | **0.047 (0.0079)** |  |  |  |  |
| EC_AN | -0.0097 (0.1072) | -0.0039 (0.2212) | **-0.0036 (0.04)** |  |  |  |
| CON | 0.0008 (0.2982) | -0.0152 (0.2735) | 0.0052 (0.4684) | **-0.0056 (0.0152)** |  |  |
| DMN | 0.0054 (0.2969) | 0 (0.2939) | 0.0054 (0.4516) | 0.0127 (0.6267) | 0 (0.5438) |  |
| eDMN | 0.0031 (0.2875) | -0.0061 (0.4537) | 0.0161 (0.3629) | -0.0144 (0.1081) | 0.0067 (0.3536) | 0.0088 (0.5713) |

(d) CURRENTLY DEPRESSED
| Functional Subnetwork | SMN | VN | EC_AN | CON | DMN | eDMN |
| --- | --- | --- | --- | --- | --- | --- |
| SMN | -0.0081 (0.2416) |  |  |  |  |  |
| VN | 0 (0.3459) | **0.0141 (0.0185)** |  |  |  |  |
| EC_AN | 0.0004 (0.1101) | 0.0072 (0.0833) | 0.0012 (0.2388) |  |  |  |
| CON | -0.0007 (0.2894) | 0.0044 (0.2416) | -0.002 (0.1833) | 0 (0.4673) |  |  |
| DMN | -0.0028 (0.6419) | 0 (0.5312) | -0.0026 (0.6756) | -0.0064 (0.1018) | 0 (0.73) |  |
| eDMN | -0.0046 (0.2894) | -0.0002 (0.0667) | 0.0035 (0.1769) | 0 (0.1757) | 0.0034 (0.6882) | 0 (0.4673) |

(e) RECURRENT DEPRESSION
| Functional Subnetwork | SMN | VN | EC_AN | CON | DMN | eDMN |
| --- | --- | --- | --- | --- | --- | --- |
| SMN | **0.0058 (0.0094)** |  |  |  |  |  |
| VN | 0 (0.2293) | **0.0062 (0.0341)** |  |  |  |  |
| EC_AN | -0.0087 (0.1756) | -0.0063 (0.0801) | -0.0059 (0.1276) |  |  |  |
| CON | 0.002 (0.1171) | -0.0044 (0.6241) | -0.0035 (0.3898) | 0.0124 (0.086) |  |  |
| DMN | 0.001 (0.2512) | 0 (0.4158) | -0.0056 (0.468) | 0 (0.2034) | 0 (0.6463) |  |
| eDMN | 0.0038 (0.1171) | 0.0057 (0.6241) | -0.0037 (0.5942) | 0.0048 (0.312) | 0.008 (0.2052) | -0.0059 (0.72) |

### Significant feature occurrence in subnetworks for different MDD phenotypes

Table 3 - 6 show the results from the binomial test and the normalised aggregated predictive coefficients for the subnetwork functional connectivity.

For Corr without the CTQ threshold, the binomial test showed that there were significantly more edges selected as important features from the connections within the visual network (VN) in all MDD phenotypes except for the Depression medicated (*p* = 0.0002 *−* 0.0341), and they were positively predictive for MDD (*β*_*norm*_ = 0.006 *−* 0.047). Significantly more edges were selected as important features from the connections within the sensorimotor (SMN) and were positively predictive for Depression Medicated and Probable Moderate/Severe Depression phenotypes (*p* = 0.0077 *−* 0.0485, *β*_*norm*_ = 0.005 *−* 0.011). On the other hand, significantly more important edges were selected from the connections within the cingulo-opercular network (CON) and were negatively predictive for Ever Depressed and Currently Depressed phenotypes (*p* = 0.0152 *−* 0.0375, *β*_*norm*_ = *−*0.007 *− −*0.006), see Table 3. With the CTQ threshold, significantly more edges were selected as important features from the connections within the SMN and also between SMN and other subnetworks for Ever Depressed, Currently Depressed, Recurrent Depression and Probable Moderate/Severe Depression phenotypes (*p* = 0.0120 *−* 0.0473). General predictive directions for MDD were positive for connections between SMN and default mode network (DMN), between SMN and extended default more network (eDMN) and within SMN (*β*_*norm*_ = 0.001 *−* 0.026), and negative for connections between SMN and executive control and attention network (EC_AN), and between SMN and VN (*β*_*norm*_ = *−*0.005 *− −*0.020). Significantly more important edges were selected as important features from the connections within the VN and were positively predictive Ever Severely Depressed, Currently Depressed and Recurrent Depression phenotypes (*p* = 0.001 *−* 0.0417, *β*_*norm*_ = 0.013 *−* 0.072), see Table 4.

**Table 4.** Normalised predictive coefficient between or within subnetworks for the six MDD classification with CTQ threshold at 0.4 based on correlation functional connectomes. The *p*-values from binomial test were in the brackets, red cell with bold text= significantly fewer edges, green cell with bold text = significantly more edges. Sensorimotor network = SMN, visual network = VN, executive control and attention network = EC_AN, cingulo-opercular network = CON, default mode network = DMN and extended default mode network = eDMN

| Functional Subnetwork | SMN | VN | EC_AN | CON | DMN | eDMN |
| --- | --- | --- | --- | --- | --- | --- |
| SMN | -0.0082 (0.1777) |  |  |  |  |  |
| VN | -0.0093 (0.0777) | 0 (0.723) |  |  |  |  |
| EC_AN | -0.0068 (0.5843) | -0.01 (0.115) | 0.011 (0.45) |  |  |  |
| CON | -0.0034 (0.3317) | -0.0031 (0.5708) | -0.0078 (0.4968) | 0.004 (0.6751) |  |  |
| DMN | 0.0023 (0.1282) | 0 (0.3755) | 0.0029 (0.1746) | 0.0031 (0.0965) | 0 (0.6143) |  |
| eDMN | 0.0029 (0.3317) | 0.0088 (0.1777) | -0.0027 (0.3204) | -0.0052 (0.5095) | 0.0032 (0.4751) | -0.009 (0.1105) |

(a) DEPRESSION MEDICATED
| Functional Subnetwork | SMN | VN | EC_AN | CON | DMN | eDMN |
| --- | --- | --- | --- | --- | --- | --- |
| SMN | -0.0064 (0.0958) |  |  |  |  |  |
| VN | 0.001 (0.4923) | 0.0077 (0.1017) |  |  |  |  |
| EC_AN | **-0.0051 (0.0461)** | 0.0017 (0.3689) | 0.0061 (0.3651) |  |  |  |
| CON | -0.0056 (0.3566) | 0.0062 (0.0958) | **-0.0017 (0.0422)** | **-0.0051 (0.0452)** |  |  |
| DMN | 0.0001 (0.368) | 0 (0.1954) | **-0.0012 (0.0333)** | 0.0004 (0.44) | -0.0041 (0.1938) |  |
| eDMN | 0.0027 (0.1114) | 0.0015 (0.2245) | 0.0073 (0.3322) | -0.0002 (0.1815) | 0.0014 (0.332) | 0.0031 (0.4194) |

(b) EVER DEPRESSED
| Functional Subnetwork | SMN | VN | EC_AN | CON | DMN | eDMN |
| --- | --- | --- | --- | --- | --- | --- |
| SMN | 0.0054 (0.5708) |  |  |  |  |  |
| VN | -0.0102 (0.2191) | **0.0129 (0.0417)** |  |  |  |  |
| EC_AN | -0.0068 (0.2558) | 0.0009 (0.5179) | 0.005 (0.0914) |  |  |  |
| CON | 0.0055 (0.164) | **-0.0248 (0.0032)** | -0.0017 (0.3204) | 0.0101 (0.6751) |  |  |
| DMN | 0.0036 (0.4173) | 0.0087 (0.253) | -0.0123 (0.197) | 0.0081 (0.2535) | 0.0107 (0.0849) |  |
| eDMN | -0.0012 (0.4373) | -0.0044 (0.5708) | 0.005 (0.4968) | -0.0001 (0.5095) | 0.0038 (0.4751) | 0.0051 (0.6751) |

(c) EVER SEVERELY DEPRESSED
| Functional Subnetwork | SMN | VN | EC_AN | CON | DMN | eDMN |
| --- | --- | --- | --- | --- | --- | --- |
| SMN | **0.0145 (0.0472)** |  |  |  |  |  |
| VN | **-0.0194 (0.0221)** | **0.0718 (0.001)** |  |  |  |  |
| EC_AN | 0 (0.158) | **-0.0012 (0.0232)** | -0.0158 (0.2194) |  |  |  |
| CON | 0.0017 (0.3589) | 0.0009 (0.315) | -0.0067 (0.5554) | 0.0079 (0.5305) |  |  |
| DMN | **0.0256 (0.0473)** | -0.0074 (0.6211) | -0.004 (0.3876) | 0 (0.09) | 0 (0.5169) |  |
| eDMN | 0.0168 (0.2002) | -0.0121 (0.315) | **0.0102 (0.0422)** | -0.0031 (0.124) | -0.0047 (0.3108) | 0.0068 (0.5305) |

(d) CURRENTLY DEPRESSED
| Functional Subnetwork | SMN | VN | EC_AN | CON | DMN | eDMN |
| --- | --- | --- | --- | --- | --- | --- |
| SMN | -0.0184 (0.2303) |  |  |  |  |  |
| VN | -0.0097 (0.732) | **0.034 (0.0174)** |  |  |  |  |
| EC_AN | **-0.0166 (0.012)** | 0 (0.1899) | 0.0222 (0.5588) |  |  |  |
| CON | 0.0075 (0.4499) | **0.0318 (0.0123)** | -0.0059 (0.1979) | -0.0181 (0.1641) |  |  |
| DMN | **0.0097 (0.0316)** | 0 (0.5421) | -0.0215 (0.1258) | -0.0097 (0.7018) | 0 (0.7374) |  |
| eDMN | -0.026 (0.2726) | 0 (0.3973) | 0.0076 (0.3311) | 0 (0.1859) | 0 (0.3292) | 0 (0.4789) |

(e) RECURRENT DEPRESSION
| Functional Subnetwork | SMN | VN | EC_AN | CON | DMN | eDMN |
| --- | --- | --- | --- | --- | --- | --- |
| SMN | **0.0055 (0.0291)** |  |  |  |  |  |
| VN | 0 (0.142) | 0 (0.6804) |  |  |  |  |
| EC_AN | **-0.0052 (0.0391)** | -0.0036 (0.401) | **-0.0026 (0.0383)** |  |  |  |
| CON | -0.0019 (0.3208) | -0.0017 (0.4814) | -0.0026 (0.3412) | 0.0025 (0.5965) |  |  |
| DMN | 0.001 (0.1983) | 0.0016 (0.6797) | -0.0011 (0.1095) | 0.0014 (0.3815) | 0 (0.5606) |  |
| eDMN | **0.0013 (0.0211)** | -0.0035 (0.2488) | 0.0003 (0.5197) | 0.0005 (0.3925) | 0.0013 (0.153) | 0 (0.2469) |

(f) PROBABLE MODERATE/SEVERE DEPRESSION

For pCorr without the CTQ threshold, the binomial test showed that there were significantly more edges selected as important features from the connections within the SMN in all MDD phenotypes except for the Depression medicated and Currently Depressed phenotypes (*p* = 0.0003 *−* 0.0490), and they were positively predictive for MDD (*β*_*norm*_ = 0.001 *−* 0.007). Significantly more edges were selected as important features from the connections within the VN and were positively predictive for Ever Severely Depressed and Currently Depressed phenotypes (*p* = 0.0026 *−* 0.0045, *β*_*norm*_ = 0.030 *−* 0.052), see Table 5. With the CTQ threshold, the important subnetwork connections as well as the sign of the predictive coefficients were quite different across the MDD phenotypes. For instance, according to the classification models, connections within the VN were important for predicting Ever Depressed and Currently Depressed phenotypes (*p* = 0.0031 *−* 0.0142). They were positively predictive for Currently Depressed (*β*_*norm*_ = 0.050) but negatively predictive for Ever Depressed (*β*_*norm*_ = *−*0.030). The connections in SMN - EC_AN were important for predicting Depression Medicated and Recurrent Depression phenotypes (*p* = 0.0008 *−* 0.0315), where they were negatively predictive for Depression Medicated (*β*_*norm*_ = *−*0.014) but positively predictive for Recurrent Depression (*β*_*norm*_ = 0.010), see Table 6.

**Table 5.** Normalised predictive coefficient between or within subnetworks for the six MDD classification without CTQ threshold based on partial correlation functional connectomes. The *p*-values from binomial test were in the brackets, red cell with bold text = significantly fewer edges, green cell with bold text = significantly more edges. Sensorimotor network = SMN, visual network = VN, executive control and attention network = EC_AN, cingulo-opercular network = CON, default mode network = DMN and extended default mode network = eDMN

| Functional Subnetwork | SMN | VN | EC_AN | CON | DMN | eDMN |
| --- | --- | --- | --- | --- | --- | --- |
| SMN | **0.0074 (0.0096)** |  |  |  |  |  |
| VN | -0.0002 (0.6711) | 0 (0.6667) |  |  |  |  |
| EC_AN | -0.0126 (0.1936) | 0.0069 (0.2212) | -0.0024 (0.3562) |  |  |  |
| CON | 0.0007 (0.1549) | -0.004 (0.4537) | -0.0028 (0.1544) | 0 (0.2294) |  |  |
| DMN | -0.0085 (0.0948) | 0.0063 (0.6575) | 0.0028 (0.0933) | -0.0051 (0.1741) | 0 (0.5438) |  |
| eDMN | 0.0027 (0.1549) | 0.0142 (0.1087) | 0.0013 (0.4684) | -0.0003 (0.3577) | -0.003 (0.1741) | 0 (0.2294) |

(a) DEPRESSION MEDICATED
| Functional Subnetwork | SMN | VN | EC_AN | CON | DMN | eDMN |
| --- | --- | --- | --- | --- | --- | --- |
| SMN | **0.0018 (0.0003)** |  |  |  |  |  |
| VN | 0.0012 (0.329) | 0.0035 (0.1146) |  |  |  |  |
| EC_AN | 0.003 (0.2131) | 0.0011 (0.3181) | 0.0004 (0.2965) |  |  |  |
| CON | -0.0026 (0.5777) | 0.0001 (0.5146) | -0.0017 (0.2604) | **0.0053 (0.0184)** |  |  |
| DMN | -0.0002 (0.3199) | 0 (0.1729) | -0.0004 (0.5156) | 0.0014 (0.6182) | -0.0062 (0.0569) |  |
| eDMN | -0.0002 (0.3977) | 0.0004 (0.5146) | 0.0059 (0.3217) | -0.0024 (0.3036) | 0.002 (0.3908) | 0.0033 (0.6535) |

(b) EVER DEPRESSED
| Functional Subnetwork | SMN | VN | EC_AN | CON | DMN | eDMN |
| --- | --- | --- | --- | --- | --- | --- |
| SMN | -0.0091 (0.5604) |  |  |  |  |  |
| VN | -0.0106 (0.0825) | **0.0516 (0.0045)** |  |  |  |  |
| EC_AN | 0.0057 (0.3808) | 0.013 (0.5041) | -0.0268 (0.0976) |  |  |  |
| CON | -0.0106 (0.4231) | 0.0098 (0.5604) | -0.0242 (0.3381) | 0.0041 (0.1157) |  |  |
| DMN | **0.0248 (0.0137)** | 0.0101 (0.2605) | -0.0176 (0.6085) | 0 (0.1628) | 0.0196 (0.0879) |  |
| eDMN | -0.0072 (0.1944) | -0.009 (0.5604) | -0.0007 (0.4781) | 0.014 (0.1275) | 0.0068 (0.4639) | 0 (0.3004) |

(c) EVER SEVERELY DEPRESSED
| Functional Subnetwork | SMN | VN | EC_AN | CON | DMN | eDMN |
| --- | --- | --- | --- | --- | --- | --- |
| SMN | 0.0128 (0.1119) |  |  |  |  |  |
| VN | -0.0202 (0.1536) | **0.0299 (0.0026)** |  |  |  |  |
| EC_AN | -0.0017 (0.3044) | -0.0158 (0.0829) | 0.0013 (0.4767) |  |  |  |
| CON | -0.0102 (0.5996) | 0 (0.0754) | 0.0131 (0.2958) | -0.0171 (0.6667) |  |  |
| DMN | 0.001 (0.3354) | 0 (0.1801) | **0.0038 (0.0248)** | 0.0147 (0.0857) | 0 (0.4262) |  |
| eDMN | 0.0026 (0.1096) | -0.0059 (0.2734) | 0.0098 (0.1115) | **-0.0221 (0.0395)** | -0.0041 (0.6348) | 0.0197 (0.0537) |

(d) CURRENTLY DEPRESSED
| Functional Subnetwork | SMN | VN | EC_AN | CON | DMN | eDMN |
| --- | --- | --- | --- | --- | --- | --- |
| SMN | **0.0013 (0.049)** |  |  |  |  |  |
| VN | -0.0009 (0.4356) | 0.0036 (0.1168) |  |  |  |  |
| EC_AN | **6.85e-6 (0.0228)** | 0.0011 (0.3044) | -0.0102 (0.5941) |  |  |  |
| CON | -0.0062 (0.5668) | -0.0008 (0.5069) | 0.0013 (0.5288) | -0.0047 (0.6468) |  |  |
| DMN | 0.002 (0.2755) | 0 (0.1694) | 0 (0.1787) | -0.0021 (0.1707) | -0.0039 (0.2195) |  |
| eDMN | -0.0012 (0.2751) | 0.0027 (0.2707) | 0.0016 (0.5288) | **0.0027 (0.0477)** | 0.0013 (0.1707) | 0.0049 (0.6468) |

(e) RECURRENT DEPRESSION
| Functional Subnetwork | SMN | VN | EC_AN | CON | DMN | eDMN |
| --- | --- | --- | --- | --- | --- | --- |
| SMN | **0.002 (0.022)** |  |  |  |  |  |
| VN | 0.0007 (0.1029) | 0 (0.699) |  |  |  |  |
| EC_AN | -0.0045 (0.3444) | 0.0005 (0.4508) | -0.0239 (0.2553) |  |  |  |
| CON | 0.003 (0.1608) | 0.0012 (0.2166) | 0.0014 (0.1376) | -0.0008 (0.1374) |  |  |
| DMN | 0.0039 (0.3632) | -0.0051 (0.7095) | -0.0037 (0.126) | 0.0066 (0.3032) | -0.0092 (0.1004) |  |
| eDMN | **0 (0.0364)** | 0 (0.1958) | **0.01 (0.0295)** | -0.0033 (0.2076) | 0 (0.1404) | 0.0064 (0.631) |

(f) PROBABLE MODERATE/SEVERE DEPRESSION

**Table 6.**
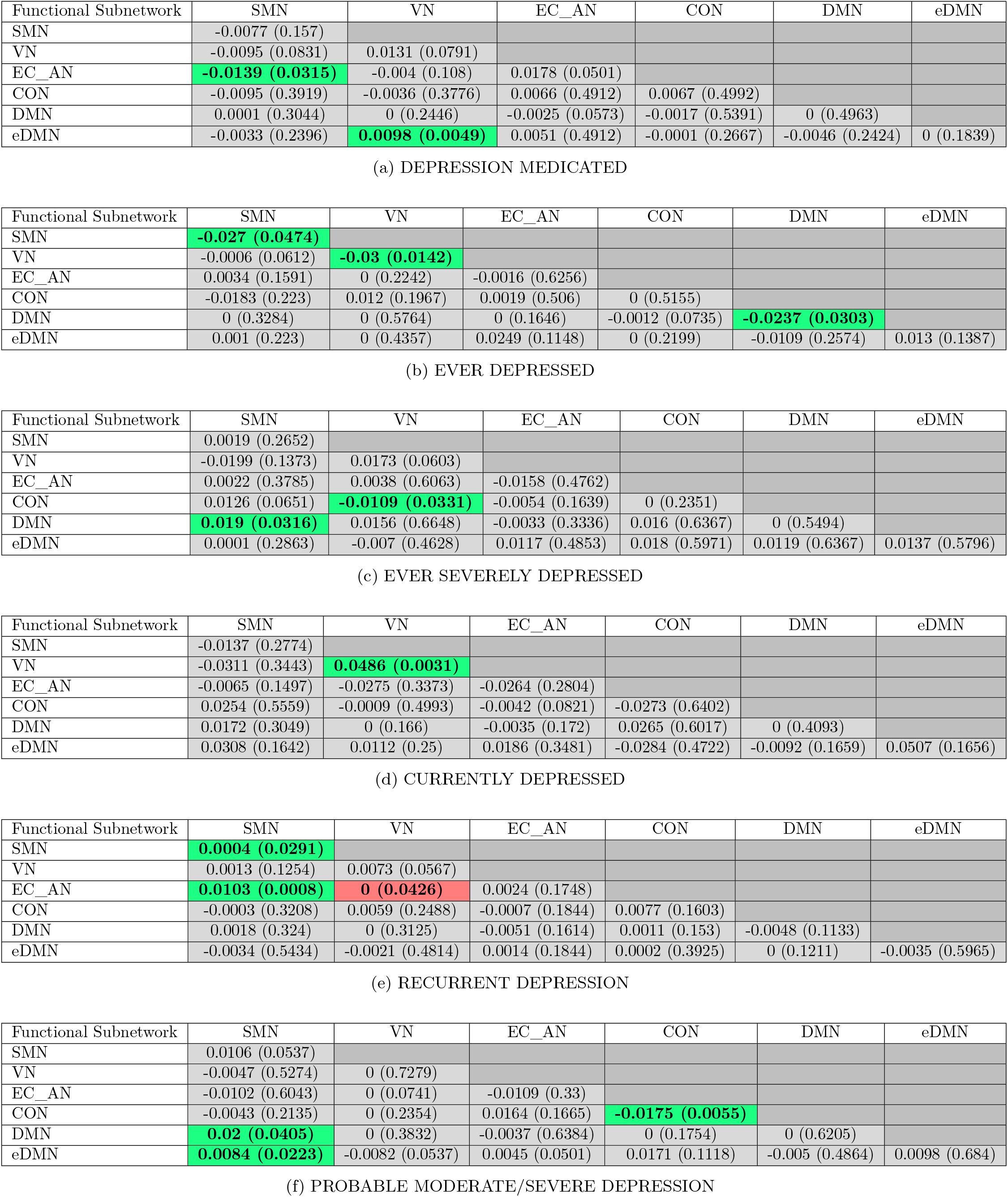
Normalised predictive coefficient between or within subnetworks for the six MDD classification with CTQ threshold at 0.4 based on partial correlation functional connectomes. The *p*-values from binomial test were in the brackets, red cell with bold text = significantly fewer edges, green cell with bold text = significantly more edges. Sensorimotor network = SMN, visual network = VN, executive control and attention network = EC_AN, cingulo-opercular network = CON, default mode network = DMN and extended default mode network = eDMN

### Comparing feature occurrence in subnetworks between models with and without the CTQ threshold for currently depressed phenotype

#### Correlation matrices

The results showed that significantly more edges were being selected from connections within SMN, within VN, between SMN and VN, and between VN and EC_AN for the model with CTQ threshold (CTQmodel). This was similar to the results from the model without the CTQ threshold (NoCTQmodel), where there were more edges being selected from connections within SMN, within VN, between SMN and VN, and between VN and EC_AN (although it did not reach statistical significance for connections within SMN, between SMN and VN, and between VN and EC_AN). The signs of the normalised aggregated predictive coefficients of these four connections of the NoCTQmodel matched with that of the CTQmodel. There were significantly fewer edges selected from connections within the EC_AN for the NoCTQmodel, which was similar to the CTQmodel although it was not significant in the CTQmodel, and the sign of the predictive coefficient match.

As for the differences, results showed that the connections between the DMN and SMN were significantly more frequently selected as important features for the CTQmodel while the same connections were less frequently selected for the NoCTQmodel. The connections between the EC_AN and eDMN were significantly less frequently selected as important features for the CTQmodel while the same connections were more frequently selected for the NoCTQmodel. The connections within CON were significantly more frequently selected as important features for the NoCTQmodel while the same connections were less frequently selected for the CTQmodel, the sign of the predictive coefficients were opposite to each other, see Figure 4a - 4b.

**Figure 4.**
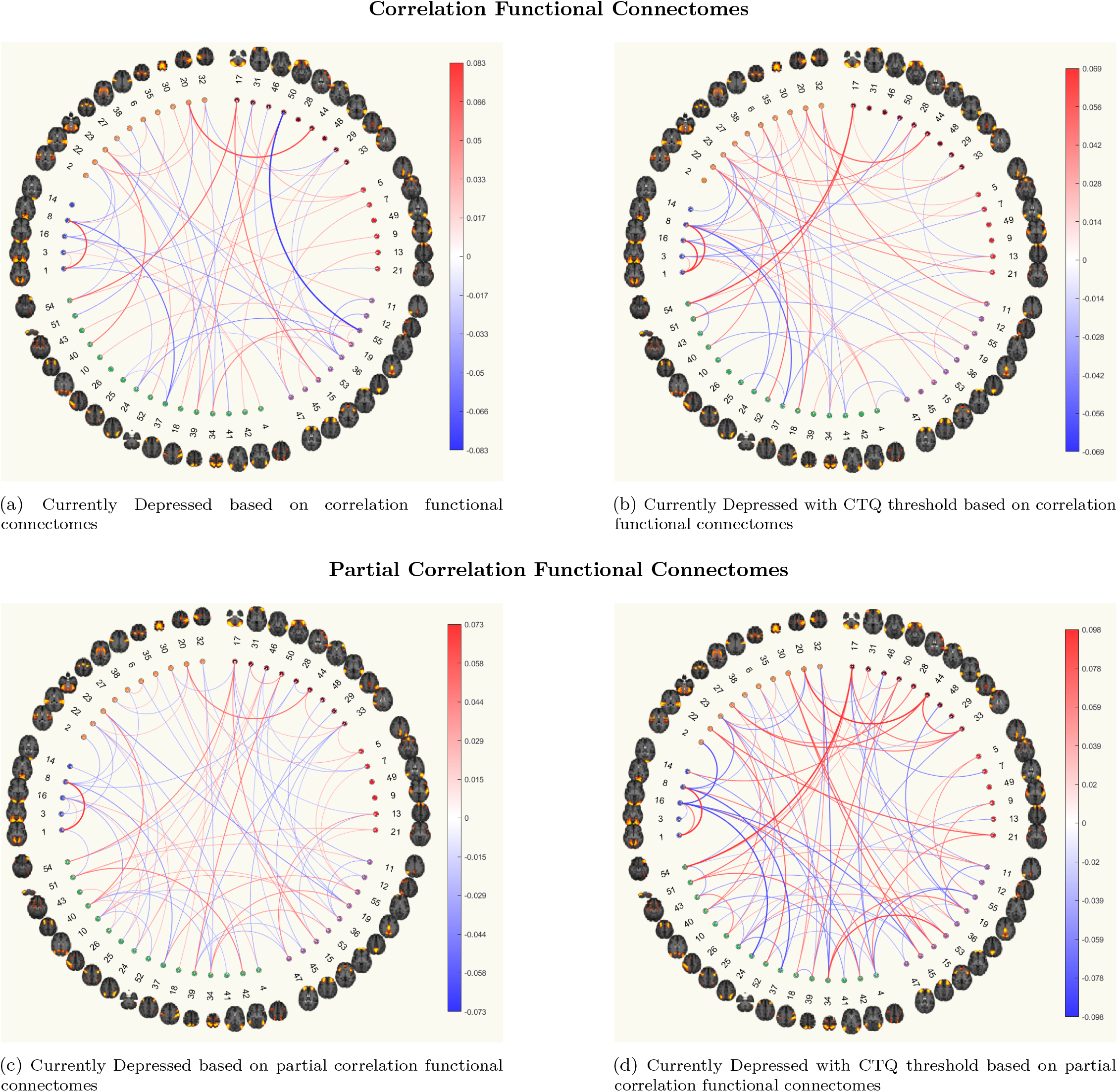
The circular plots showing important features for predicting Currently Depressed phenotype with and without the CTQ threshold based on functional connectomes. The ordering of the 55 good ICA nodes is the same as that from the UKB functional network interactive visualisation, where the coloured clusters approximately maps to SMN (orange), VN (blue), EC_AN (green), CON (purple), DMN (red) and eDMN (brown). Details of method for identifying important features are presented in Methods section.

#### Partial correlation matrices

For partial correlation matrices, there were significantly more edges selected as important features from the connections within VN for both the NoCTQmodel and the CTQmodel as mentioned in the above section. In addition, we saw that the connections between the DMN and EC_AN were significantly less frequently selected as important feature for the NoCTQmodel while in the CTQmodel, it was not significant and the predictive coefficient’s sign was opposite to that of the NoCTQmodel, see Figure 4c - 4d.

### Overall findings in functional connectomes

The important features indicated by the models were largely different across different MDD phenotypes. The connections with SMN and VN, whether it was within subnetwork connections or with other subnetworks, were selected more frequently as important features for predicting MDD phenotypes, while past studies mainly found aberrant functional connectivity within the DMN in depressed patients (Kaiser et al., 2015; Mulders et al., 2015). We are aware of the fact that these results are presented in the context of performing ML classifications, and the important features being selected here only implied that they were the edges consistently having top rankings in terms of coefficient magnitudes across folds. Therefore, the results presented in the current study does not necessarily contradict the previously reported abnormal functional connectivity within DMN in MDD patients - shared covariance for functional connectivity in these networks may well have been more consistently and strongly represented (independently of all other features) in other edges as selected by the ML classifiers across the nested cross-validation folds. As far as the effect of CTQ thresholding is concerned, in addition to the low Jaccard index seen in Table 2a and Table 2b, some of the overlapping edges had coefficients with opposing associations. The models with CTQ thresholding usually had more important edges selected, which indicated that the magnitudes and direction of the models’ coefficients were more consistent for classifying MDD with a CTQ threshold, giving partial evidence to the claim that MDD with early adversity features forms a more homogeneous subgroup.

## Discussion

In the current study, we applied logistic ridge regression to classify depression phenotypes based on functional and structural connectomes, where we ensured the 1-to-1 case-control matching and correction for confounders throughout the classification analyses for all the different MDD phenotypes. In general, the MDD classification test accuracies based on functional connectomes were higher than those based on structural connectomes, though accuracy was not > 62% for any model. The classification test accuracies for Currently Depressed phenotype were the highest among all the MDD phenotypes. We also found that adding childhood adversity information to the depression case criterion benefited model accuracy for some of the models, namely Currently Depressed phenotype based on both functional and structural connectomes except for ICVF, Ever Severely Depressed phenotype based on functional connectomes but not for structural connectomes, and Probable Moderate/Severe Depression phenotype based on MD, FA, ICVF and ISOVF.

We found that ML models based on SC connectomes exhibited the lowest accuracies among all the structural network weightings. Previous research usually found SC (or edge density, a variant of SC) had stronger associations with other clinical and behavioural phenotypes (Oestreich et al., 2019). Buchanan et al. (2020) have commented that the confounding effect of age, sex and head size on SC weightings could be considerably larger than on other types of network weightings, which may have contributed to the superior predictive performances of SC when predicting psychiatric disorders shown in previous studies (Payabvash et al., 2019; Raji et al., 2020). In the current study, we chose healthy controls with age, sex and ICV matched (or closest to) with the cases in order to minimize the confounding effect and this could be the reason for having results inconsistent with the previous literature. In addition, a similar phenomenon was seen in our previous paper where model classification performances on general cognitive function and general psychopathology were comparable for all the structural connectivity modalities when age and sex were added to the models (Yeung et al., 2022).

In terms of MDD phenotyping, we found that the more severe phenotype usually got higher classification accuracies. It is possible that there is less heterogeneity in the more severe MDD group and therefore easier to classify. However, smaller sample size could also have contributed to the boost in classification accuracies. It is worth highlighting that we saw a moderate-to-strong negative correlations between test accuracies and sample size, which is consistent with the results in Stolicyn et al. (2020).

When considering the important network features, we found that less than 10 percent of the total number of edges were considered as important features for predicting any type of depression phenotype based on any connectome modality in this study. We also found that the set of important features were different between models with and without CTQ threshold. This may have partly demonstrated the neurobiological differences between MDD patients with and without childhood adversity, which was reported in previous studies (Fadel et al., 2021; Luo et al., 2022a) and we showed this in a much larger dataset than in previous studies. However, it is also possible that by adding an addition CTQ thresholding constraint on the MDD cases, we are essentially defining a more severe type of MDD based on the given MDD phenotype (Huh et al., 2017). This may explain why there is higher similarity, higher Jaccard index, between the feature sets identified by the CTQmodels and the NoCTQmodel for the more severe MDD phenotypes.

For resting-state functional connectivity, in contrast to most previous studies where they either found significant hyperconnectivity or hypoconnectivity within DMN in depressed patients (Hamani et al., 2011; Kaiser et al., 2015; Sheline et al., 2010; Yan et al., 2019), we found that the DMN was not often selected as the hub of important connections for classifying MDD, and sometimes even less than random chance. The connections within the DMN are not selected significantly more often than random chance in any of the MDD phenotypes based on either pCorr or Corr, except for the Ever Depressed phenotype with CTQ threshold of 0.4 based on pCorr where connections within the DMN are negatively predictive for MDD. On the other hand, we found that the connections from as well as within SMN and VN were more frequently selected as important features for predicting MDD than other subnetworks. For the models which indicated SMN and VN as important hubs for MDD classification, the connectivity within SMN and VN were mostly positive predictive for MDD. The fact connections from SMN and VN were more frequently selected by the models as important features for classifying MDD across different MDD phenotypes possibly suggest that differences in networks involved in processing of sensory information may be a more stable neuroimaging marker for the purposes of MDD prediction out-of-sample (Javaheripour et al., 2021).

There are a few limitations to this study. First, different atlas for parcellation of the nodes for the structural and functional connectomes at the preprocessing step, where group Independent Component Analysis (group-ICA) was used for functional connectomes and the Desikan-Killiany atlas was used for the structural connectomes. This partly limits direct neuroanatomical comparisons of selected features for structural and functional connectivity and interpretations on the added value of combining structural and functional connectivity in MDD classifications. Second, representing functional connectivity in the form of one adjacency matrix per person may not be the most optimal way. There are studies with smaller data sets which use functional connectomes in time series form and applied the hybrid model, combining Graph Convolutional Neural Network (GCNN) and Long-Short Term Memory (LSTM) Network for predicting clinical phenotypes. They reported that the hybrid models performed better at classifying autistic patients compared to other ML models (Masood and Kashef, 2022; Wang et al., 2021). Similarly, structural network measures are known to be sensitive to the network construction methodology (Qi et al., 2015). Therefore, different structural findings might be obtained with connectome methods different to those applied here. Fourth, the CTQ items from the UKB have an abbreviated scale and are potentially confounded by current mood, potentially introducing bias (Madden et al., 2022). In addition, different types of trauma are likely to have differential effect on MDD individuals. Certain types of childhood trauma have been suggested to be more associated with symptom severity in MDD patients, while different types of trauma have been linked to differential alteration in MDD brain functional connectivity (Fadel et al., 2021; Negele et al., 2015). A more comprehensive and detailed CTQ is needed to thoroughly investigate the effect of childhood adversity on depression. Fourth, the UKB consists of healthier, wealthier and older individuals (Fry et al., 2017) and only a small portion of the MDD cases meet the criteria for current depression. Although it was found that the currently depressed phenotype achieved the best classification accuracies, the small sample size could have been the reason for the high accuracy, as shown in the Results. Similar results were also shown in previous study (Stolicyn et al., 2020). These results provide the basis for further replication and refinement in other community samples and in clinical cohorts.

In conclusion, this study reports a comprehensive data-driven MDD classification analysis, classifying six MHQ-derived MDD phenotypes based on a wide range of functional and structural brain connectivity measures, on a large community sample (UKB). The results indicated a positive relationship between depression severity and classification accuracy. Our findings also suggested that SMN and VN are robust and important biomarkers of MDD. The model for Currently Depressed phenotype based on pCorr achieved the highest accuracy of 61.06%, and the accuracy for classifying this phenotype with a CTQ threshold was 4.68% higher. The set of important features selected for models with and without CTQ threshold were largely different. Connections from SMN and VN were more frequently selected for classifying MDD across different MDD phenotypes rather than DMN and EC_AN. The robust functional connectivity features for MDD classification identified in the current study were in line with the recent imaging studies in MDD. This provided the basis to verify and build on the idea of having sensory related subnetworks as one of the possible robust biomarkers for MDD.

## Data Availability

The UK Biobank's Access Procedures stipulate that participant data can only be made available to approved researchers. Therefore, the data used in this study cannot be made available for public access.

## Acknowledgements

This study is supported by Wellcome Trust awards (References 104036/Z/14/Z; 220857/Z/20/Z), and was also supported by National Institutes of Health (NIH) research grant R01AG054628 which supported CRB, EMTD, MEB and SRC. The research was conducted using the UK Biobank resource, with approved project number 10279 and 4844. Structural brain imaging data from UK Biobank was processed using facilities within the Lothian Birth Cohort group at the University of Edinburgh, which is supported by Age UK (as The Disconnected Mind project), the Medical Research Council (MR/R024065/1), and the University of Edinburgh. This work has made use of the resources provided by the Edinburgh Compute and Data Facility (ECDF). The Population Research Center (PRC) and Center on Aging and Population Sciences (CAPS) at The University of Texas at Austin are supported by National Institutes of Health (NIH) grants P2CHD042849 and P30AG066614, respectively. HWY is supported by the endowments to the Division of Division of Psychiatry, University of Edinburgh. GT is supported by supported by the Agency of Science, Technology and Research (A*STAR) in Singapore. KMS was supported by Health Data Research UK, an initiative funded by UK Research and Innovation Councils, NIH Research (England) and the UK devolved administrations, and leading medical research charities. SRC is also supported by a Sir Henry Dale Fellowship jointly funded by the Wellcome Trust and the Royal Society (221890/Z/20/Z). AMM and HCW are additionally supported by a UKRI award (Reference MC_PC_17209).

