## Supplementary material for "Classification accuracy of structural and functional connectomes across different depressive phenotypes": NA

[1]Department of Psychiatry, University of Edinburgh, Edinburgh, United Kingdom
[2]Department of Psychology, University of Edinburgh, Edinburgh, United Kingdom
[3]Department of Psychology, University of Texas, Austin, TX, USA
[4]Population Research Center and Center on Aging and Population Sciences, University of Texas at Austin, TX, USA
[5]Centre for Clinical Brain Science, University of Edinburgh, Edinburgh, United Kingdom
[6]Centre for Genomic and Experimental Medicine, Institute of Genetics and Molecular Medicine, University of Edinburgh, Edinburgh, UK
[7]Department of Physics and Mathematics, Nottingham Trent University, Nottingham, United Kingdom
[8]Lothian Birth Cohorts, University of Edinburgh, Edinburgh, United Kingdom
[9]Scottish Imaging Network, A Platform for Scientific Excellence Collaboration (SINAPSE), Edinburgh, United Kingdom
[*]These authors share joint senior authorship

November 22, 2022

### A Materials and Methods

#### A.1 Depression Definitions

The first five definitions (Depression Medicated, Ever Depressed, Ever Severely Depressed, Currently Depressed, Recurrent Depression) were presented in Davis et al. (2020) in details, and the R code is available on (Coleman and Davis, 2019). The detail description of the last MDD phenotype, probable moderate/severe depression, was presented in Smith et al. (2013). For clarification, the brief description and the code for the six MDD phenotypes used in this study were shown below.

| MDD phenotypes | Description | Field and code |
|---|---|---|
| Depression Medicated | Taken at least one of the medication listed on the antidepressants list | Please refer to the R code for the antidepressant fields |
| Ever Depressed | At least one core symptom of depression, most or all of the day on most or all days for a two week period, with at least five depressive symptoms that represent a change from usual occurring over the same time-scale, with some or a lot of impairment. | Persistent sadness (20446) = Yes OR Loss of interest (20441) = Yes<br>AND<br>How much of day (20436) = Most of day or All day long<br>AND<br>Did you feel this way (20439) = Almost every day or Every day<br>AND<br>Impairment (20440) = Somewhat or A lot<br>AND |

| MDD phenotypes | Description | Field and code |
|---|---|---|
| | | Total number of symptoms endorsed (core and others) $\geq 5$<br>Persistent sadness (core) 20446; Loss of interest (core) 20441; Tired or low energy 20449; Gain or loss of weight 20536 = Gain, Loss or Gain and loss; Sleep change 20532; Trouble concentrating 20435; Feeling worthless 20450; Thinking about death 20437 |
| Ever Severely Depressed | | {Ever Depressed}<br>AND<br>CIDI Severity Score = 8 |
| Currently Depressed | PHQ+ve and CIDI+ve Reports symptoms in the last two weeks that have bothered them. Current depression is indicated by five or more items marked to bother at or above a certain intensity: "more than half of days" for first eight items, "some days" for last item. | {Ever Depressed}<br>AND<br>Total symptoms endorsed as occurring more than half days (or some or more days for last item) $\geq 5$<br>Little interest or pleasure in doing things 20514, Feeling down, depressed, or hopeless 20510, Trouble sleeping 20517, Feeling tired 20519, Poor appetite or overeating 20511, Feeling bad about yourself 20507, Trouble concentrating 20508, Moving or speaking slowly or fidgety or restless 20518, Thoughts that you would be better off dead 20513 |
| Recurrent Depression | | {Ever Depressed}<br>AND<br>Number of episodes (20442) > 1 or -999 (too many to count)<br>AND<br>NOT case bipolar type I<br><br>Excluded if number of episodes missing or bipolar state missing |

| MDD phenotypes | Description | Field and code |
| --- | --- | --- |
| Probable Moderate/Severe Depression | Either satisfying the criteria for Probable recurrent major depression (moderate) or Probable recurrent major depression (severe) described in Smith et al. (2013). | Probable recurrent major depression (moderate):<br>EITHER:<br>4598 Ever depressed/down for a whole week, plus<br>4609 At least two weeks duration, plus<br>4620 At least two episodes, plus<br>2090 Ever seen a GP (but not a psychiatrist) for nerves, anxiety, depression<br>OR:<br>4631 Ever anhedonic (unenthusiasm / uninterest) for a whole week, plus<br>5375 At least two weeks, plus<br>5386 At least two episodes, plus<br>2090 Ever seen a GP (but not a psychiatrist) for nerves, anxiety, depression<br><br>Probable recurrent major depression (severe):<br>EITHER:<br>4598 Ever depressed/down for a whole week, plus<br>4609 At least two weeks duration, plus<br>4620 At least two episodes, plus<br>2100 Ever seen a psychiatrist for nerves, anxiety, depression<br>OR:<br>4631 Ever anhedonic (unenthusiasm / uninterest) for a whole week, plus<br>5375 At least two weeks, plus<br>5386 At least two episodes, plus<br>2100 Ever seen a psychiatrist for nerves, anxiety, depression |

#### A.2 Childhood Trauma Questionnaire

The UK Biobank Childhood Trauma Questionnaire items are listed in the following:

1. Felt loved as a child (inverse scored)
2. Someone to take me to the doctor as a child (inverse scored)
3. Sexually molested as a child
4. Physically abused by family as a child
5. Felt hated by family member as a child

which they correspond to emotional neglect, physical neglect, sexual abuse, physical abuse and emotional abuse respectively. We converted the responses to a scoring system of 0-to-4 to represent the severity (ranging from 'never true' to 'very often true') of trauma for each of the items. As for two of the positive items, it was score in an inverse way to capture trauma.

#### A.3 Jaccard Index

Jaccard index is used to measure the similarity between two set of elements, and the similarity between set $A$ and $B$ is given by:

$$Jaccard(A, B) = \frac{|A \cap B|}{|A \cup B|} \tag{A.1}$$

which is the size of intersection divided by the size of union.

#### A.4 Predictive coefficients normalisation by subnetwork size

A straight forward aggregation of predictive coefficient of the robust edge features to subnetwork level can be computed using the following:

$$S = C^T E C \tag{A.2}$$

where $E$ is a $n$-by-$n$ predictive coefficients matrix for the robust edges ($n$ is the number of nodes), $S$ is a $c$-by-$c$ matrix ($c$ is the number of subnetworks), $S_{ij}$ represents the aggregated predictive coefficient for the connectivity between subnetwork $i$ and $j$, $C$ is $n$-by-$c$ community indicator matrix, $C_{ij} = \delta(c_i, c_j)$ with $c_i$ representing the community of node $i$.

However, this is a biased measurement due to the size differences among the subnetworks. To account for this, we replace $C$ by $C'$, where

$$C'_{ij} = \frac{C_{ij}}{\sqrt{\sum_{k=1}^{n} C_{kj}}} \tag{A.3}$$

### B MDD phenotypes Classification Results

Table 2: Classification performance for different depression phenotypes (with different CTQ thresholds) based on different connectome modalities. Drug = Depression Medicated, Ever = Ever Depressed, Ever Severe = Ever Severely Depressed, Current = Currently Depressed, Recurrent = Recurrent Depression without Bipolar Disorder, MDD Narrow = Probable Moderate/Severe Depression

| MDD phenotype | Drug | Ever | Ever Severe | Current | Recurrent | MDD narrow |
|---|---|---|---|---|---|---|
| Without CTQ Threshold | 55.98 (3.26) | 54.24 (1.59) | 53.10 (3.52) | 57.44 (5.33) | 54.38 (1.88) | 55.15 (2.53) |
| CTQ Threshold at 0.2 | 55.05 (3.34) | 53.04 (1.72) | 53.17 (2.92) | 60.06 (3.21) | 52.98 (1.83) | 54.79 (3.49) |
| CTQ Threshold at 0.4 | 54.64 (3.90) | 54.39 (1.54) | 53.91 (5.48) | 60.68 (4.29) | 53.07 (1.74) | 54.26 (3.07) |
| CTQ Threshold at 0.6 | 54.88 (3.90) | 53.01 (2.84) | 54.86 (7.59) | 60.57 (5.73) | 53.70 (2.92) | 55.46 (2.38) |

(a) Classification accuracies of the test set (mean percentage with standard deviation in brackets) for different depression phenotypes (with different CTQ thresholds) based on correlations functional connectome

| MDD phenotype | Drug | Ever | Ever Severe | Current | Recurrent | MDD narrow |
|---|---|---|---|---|---|---|
| Without CTQ Threshold | 59.15 (2.73) | 54.30 (1.35) | 52.43 (2.94) | 61.06 (4.82) | 54.70 (1.51) | 53.25 (2.97) |
| CTQ Threshold at 0.2 | 58.03 (3.00) | 53.73 (1.43) | 53.49 (3.13) | 65.47 (4.18) | 52.80 (1.86) | 52.58 (3.32) |
| CTQ Threshold at 0.4 | 57.42 (2.44) | 53.55 (1.85) | 54.70 (4.01) | 65.74 (4.73) | 52.62 (2.43) | 52.04 (3.20) |
| CTQ Threshold at 0.6 | 57.73 (3.46) | 52.59 (1.64) | 53.48 (5.28) | 64.87 (6.57) | 52.30 (2.26) | 53.74 (4.21) |

(b) Classification accuracies of the test set (mean percentage with standard deviation in brackets) for different depression phenotypes (with different CTQ thresholds) based on partial correlations functional connectome

| MDD phenotype | Drug | Ever | Ever Severe | Current | Recurrent | MDD narrow |
|---|---|---|---|---|---|---|
| Without CTQ Threshold | 55.53 (3.67) | 51.74 (1.81) | 53.17 (4.67) | 56.45 (6.58) | 52.98 (2.82) | 53.61 (4.90) |
| CTQ Threshold at 0.2 | 52.63 (4.12) | 53.02 (2.09) | 53.05 (4.24) | 57.56 (10.37) | 52.98 (3.53) | 53.24 (4.92) |
| CTQ Threshold at 0.4 | 54.50 (5.66) | 53.05 (2.79) | 51.53 (5.22) | 59.57 (7.06) | 52.88 (3.00) | 55.27 (4.72) |
| CTQ Threshold at 0.6 | 55.64 (6.78) | 53.80 (3.54) | 51.83 (6.08) | 59.19 (9.10) | 50.92 (3.85) | 55.93 (5.88) |

(c) Classification accuracies of the test set (mean percentage with standard deviation in brackets) for different depression phenotypes (with different CTQ thresholds) based on mean diffusivity structural connectome

| MDD phenotype | Drug | Ever | Ever Severe | Current | Recurrent | MDD narrow |
|---|---|---|---|---|---|---|
| Without CTQ Threshold | 55.77 (3.59) | 51.18 (1.81) | 55.72 (4.04) | 58.46 (10.17) | 52.70 (2.89) | 53.81 (3.76) |
| CTQ Threshold at 0.2 | 53.85 (4.53) | 53.01 (1.82) | 52.60 (4.58) | 55.77 (8.37) | 52.56 (2.96) | 52.89 (3.35) |
| CTQ Threshold at 0.4 | 55.72 (6.52) | 53.38 (2.25) | 52.25 (6.76) | 59.44 (7.00) | 54.34 (3.25) | 54.33 (6.42) |
| CTQ Threshold at 0.6 | 53.70 (8.45) | 52.95 (3.17) | 51.11 (6.40) | 63.40 (7.73) | 52.92 (4.24) | 57.51 (7.67) |

(d) Classification accuracies of the test set (mean percentage with standard deviation in brackets) for different depression phenotypes (with different CTQ thresholds) based on fractional anisotropy structural connectome

Table 2: Classification performance for different depression phenotypes (with different CTQ thresholds) based on different connectome modalities. Drug = Depression Medicated, Ever = Ever Depressed, Ever Severe = Ever Severely Depressed, Current = Currently Depressed, Recurrent = Recurrent Depression without Bipolar Disorder, MDD Narrow = Probable Moderate/Severe Depression (Cont.)

| MDD phenotype | Drug | Ever | Ever Severe | Current | Recurrent | MDD narrow |
|---|---|---|---|---|---|---|
| Without CTQ Threshold | 52.28 (3.35) | 51.27 (2.23) | 53.32 (4.87) | 50.25 (7.69) | 51.63 (2.86) | 50.38 (3.70) |
| CTQ Threshold at 0.2 | 50.20 (4.04) | 50.17 (1.95) | 50.95 (6.18) | 50.00 (7.95) | 51.16 (4.11) | 50.27 (4.20) |
| CTQ Threshold at 0.4 | 47.89 (5.29) | 49.66 (2.83) | 51.66 (4.65) | 53.60 (8.86) | 50.46 (2.98) | 48.48 (4.16) |
| CTQ Threshold at 0.6 | 48.86 (7.21) | 49.90 (3.52) | 53.49 (7.12) | 51.76 (8.74) | 50.42 (3.65) | 47.38 (4.86) |

(e) Classification accuracies of the test set (mean percentage with standard deviation in brackets) for different depression phenotypes (with different CTQ thresholds) based on streamline count structural connectome

| MDD phenotype | Drug | Ever | Ever Severe | Current | Recurrent | MDD narrow |
|---|---|---|---|---|---|---|
| Without CTQ Threshold | 53.89 (4.78) | 51.90 (1.51) | 54.76 (4.61) | 54.27 (7.34) | 52.32 (2.70) | 50.89 (3.87) |
| CTQ Threshold at 0.2 | 53.78 (5.11) | 51.51 (2.13) | 52.25 (4.62) | 56.54 (8.52) | 53.32 (2.91) | 50.18 (4.81) |
| CTQ Threshold at 0.4 | 54.33 (4.43) | 51.06 (2.44) | 52.64 (5.04) | 58.70 (7.99) | 53.11 (2.99) | 51.26 (4.44) |
| CTQ Threshold at 0.6 | 49.44 (5.87) | 51.61 (2.81) | 54.37 (4.85) | 54.69 (8.15) | 51.75 (4.46) | 49.32 (5.65) |

(f) Classification accuracies of the test set (mean percentage with standard deviation in brackets) for different depression phenotypes (with different CTQ thresholds) based on orientation dispersion structural connectome

| MDD phenotype | Drug | Ever | Ever Severe | Current | Recurrent | MDD narrow |
|---|---|---|---|---|---|---|
| Without CTQ Threshold | 52.87 (3.57) | 51.61 (2.01) | 50.87 (4.54) | 58.73 (6.02) | 52.03 (3.43) | 51.25 (4.51) |
| CTQ Threshold at 0.2 | 52.69 (4.54) | 53.31 (2.36) | 52.02 (6.13) | 59.87 (6.28) | 53.54 (3.60) | 51.46 (5.40) |
| CTQ Threshold at 0.4 | 53.22 (6.69) | 53.93 (2.37) | 51.11 (6.12) | 62.94 (9.43) | 53.48 (3.27) | 51.52 (5.15) |
| CTQ Threshold at 0.6 | 52.23 (5.91) | 53.23 (3.64) | 47.94 (4.71) | 59.07 (6.88) | 53.39 (4.68) | 54.55 (6.15) |

(g) Classification accuracies of the test set (mean percentage with standard deviation in brackets) for different depression phenotypes (with different CTQ thresholds) based on isotropic volume fraction structural connectome

| MDD phenotype | Drug | Ever | Ever Severe | Current | Recurrent | MDD narrow |
|---|---|---|---|---|---|---|
| Without CTQ Threshold | 55.40 (3.95) | 52.99 (2.01) | 56.07 (4.60) | 60.37 (9.30) | 53.42 (2.56) | 53.99 (4.33) |
| CTQ Threshold at 0.2 | 52.49 (4.47) | 53.21 (2.51) | 55.33 (4.66) | 60.64 (10.13) | 52.63 (2.93) | 54.32 (3.77) |
| CTQ Threshold at 0.4 | 53.00 (7.13) | 53.97 (2.68) | 55.23 (6.64) | 59.13 (9.10) | 53.65 (2.95) | 56.36 (4.95) |
| CTQ Threshold at 0.6 | 55.09 (6.30) | 53.54 (3.55) | 54.37 (6.38) | 59.67 (7.05) | 53.56 (4.09) | 56.45 (7.15) |

(h) Classification accuracies of the test set (mean percentage with standard deviation in brackets) for different depression phenotypes (with different CTQ thresholds) based on intra-cellular volume fraction structural connectome

#### B.1 MDD Classification results for sex specific models based on functional conectomes

Table 3: Classification performance of sex specific model for different depression phenotypes (with different CTQ thresholds) based on functional connectomes. Drug = Depression Medicated, Ever = Ever Depressed, Ever Severe = Ever Severely Depressed, Current = Currently Depressed, Recurrent = Recurrent Depression without Bipolar Disorder, MDD Narrow = Probable Moderate/Severe Depression (Cont.)

**Female MDD Classification Models**

| Test Accuracies | Medicated | Ever | Ever Severe | Current | Recurrent | MDD narrow |
|---|---|---|---|---|---|---|
| Without CTQ Threshold | 55.49 (3.59) | 54.28 (1.78) | 54.42 (3.82) | 56.53 (7.14) | 54.34 (1.91) | 55.53 (3.04) |
| CTQ Threshold at 0.2 | 53.48 (4.27) | 53.47 (2.03) | 52.86 (5.37) | 56.46 (7.18) | 53.91 (1.76) | 55.62 (4.16) |
| CTQ Threshold at 0.4 | 54.74 (3.40) | 53.77 (2.08) | 52.98 (6.45) | 58.33 (9.12) | 53.75 (2.50) | 54.67 (4.24) |
| CTQ Threshold at 0.6 | 52.72 (4.33) | 53.55 (2.97) | 55.56 (7.85) | 57.82 (10.02) | 54.81 (3.24) | 54.97 (4.22) |

(a) Classification accuracies of the test set (mean percentage with standard deviation in brackets) for different depression phenotypes (with different CTQ thresholds) for female MDD models based on correlation functional connectomes.

| Test Accuracies | Medicated | Ever | Ever Severe | Current | Recurrent | MDD narrow |
|---|---|---|---|---|---|---|
| Without CTQ Threshold | 59.16 (3.82) | 53.34 (2.30) | 54.91 (2.77) | 58.14 (5.67) | 54.52 (2.30) | 53.06 (2.64) |
| CTQ Threshold at 0.2 | 57.68 (3.32) | 53.05 (1.76) | 53.59 (3.47) | 58.04 (7.68) | 53.68 (1.94) | 52.76 (3.83) |
| CTQ Threshold at 0.4 | 55.42 (3.71) | 53.20 (2.02) | 56.38 (4.31) | 62.16 (11.91) | 52.85 (2.25) | 51.43 (3.60) |
| CTQ Threshold at 0.6 | 54.91 (5.56) | 53.32 (3.03) | 57.69 (4.74) | 65.00 (10.27) | 52.32 (3.23) | 53.48 (4.73) |

(b) Classification accuracies of the test set (mean percentage with standard deviation in brackets) for different depression phenotypes (with different CTQ thresholds) for female MDD models based on partial correlation functional connectomes.

**Male MDD Classification Models**

| Test Accuracies | Medicated | Ever | Ever Severe | Current | Recurrent | MDD narrow |
|---|---|---|---|---|---|---|
| Without CTQ Threshold | 52.86 (5.72) | 50.72 (2.34) | 53.61 (6.31) | 57.34 (8.01) | 49.67 (2.81) | 50.70 (5.07) |
| CTQ Threshold at 0.2 | 53.36 (5.32) | 50.42 (2.39) | 53.75 (7.08) | 57.21 (7.59) | 49.69 (2.53) | 50.59 (5.38) |
| CTQ Threshold at 0.4 | 53.75 (7.76) | 49.85 (2.78) | 49.99 (8.66) | 62.05 (7.81) | 46.70 (4.23) | 49.87 (4.00) |
| CTQ Threshold at 0.6 | 55.71 (7.20) | 48.94 (2.87) | 48.38 (8.70) | 56.02 (9.95) | 45.74 (5.64) | 51.88 (5.28) |

(c) Classification accuracies of the test set (mean percentage with standard deviation in brackets) for different depression phenotypes (with different CTQ thresholds) for male MDD models based on correlation functional connectomes.

| Test Accuracies | Medicated | Ever | Ever Severe | Current | Recurrent | MDD narrow |
|---|---|---|---|---|---|---|
| Without CTQ Threshold | 55.66 (4.38) | 52.86 (2.38) | 52.15 (5.03) | 56.76 (6.78) | 50.69 (2.64) | 53.04 (3.62) |
| CTQ Threshold at 0.2 | 51.94 (6.95) | 51.43 (3.35) | 48.54 (6.23) | 61.94 (9.65) | 47.34 (3.90) | 51.92 (5.03) |
| CTQ Threshold at 0.4 | 55.83 (7.89) | 51.86 (2.47) | 48.75 (9.20) | 62.90 (8.98) | 46.55 (4.21) | 51.56 (6.74) |
| CTQ Threshold at 0.6 | 57.98 (10.28) | 50.80 (4.62) | 46.33 (10.51) | 55.65 (7.42) | 45.06 (4.04) | 51.88 (9.62) |

(d) Classification accuracies of the test set (mean percentage with standard deviation in brackets) for different depression phenotypes (with different CTQ thresholds) for male MDD models based on partial correlation functional connectomes.
